# Monitoring of SARS-CoV-2 Antibodies Using Dried Blood Spot for At-Home Collection

**DOI:** 10.1101/2021.11.19.21266532

**Authors:** Peyton K. Miesse, Bradley B. Collier, Russell P. Grant

## Abstract

The utilization of vaccines to fight the spread of SARS-CoV-2 has led to a growing need for expansive serological testing. To address this, an EUA approved immunoassay for detection of antibodies to SARS-CoV-2 in venous serum samples was investigated for use with dried blood spot (DBS) samples. Results from self-collected DBS samples demonstrated a 98.1% categorical agreement to venous serum with a correlation (R) of 0.9600 while professionally collected DBS samples demonstrated a categorical agreement of 100.0% with a correlation of 0.9888 to venous serum. Additional studies were performed to stress aspects of at-home DBS collection, including shipping stability, interference effects, and other sample-specific robustness studies. These studies demonstrated a categorical agreement of at least 95.0% and a mean bias less than ±20.0%. Furthermore, the ability to track antibody levels following vaccination with the BioNTech/Pfizer vaccine was demonstrated with self-collected DBS samples from pre-dose (Day 0) out to 19 weeks.

## 1. Introduction

Despite the implementation of several measures to slow the spread of the disease, severe acute respiratory syndrome coronavirus 2 (SARS-CoV-2) continues to be an international public health emergency due to rapid human-to-human transmission and prevalence of asymptomatic carriers (*1,2*). Initially, many countries implemented physical distancing protocols and/or lockdown restrictions (*3*). In addition, diagnostic tests were quickly developed and granted FDA emergency use authorization (EUA) in order to identify individuals with active SARS-CoV-2 infections (*4*). Although social distancing and diagnostic testing continue to be vital to ending the pandemic, the advent of SARS-CoV-2 vaccines provides a more robust means of limiting the spread of the virus (*4*). By triggering the body’s natural immune response, vaccines initiate the creation of antibodies that can neutralize the virus upon infection and ultimately reduce the severity of infections as well as transmission of the virus (*5*). Unfortunately, the lifespan of circulating SARS-CoV-2 antibodies and the requisite titer to yield protective immunity against SARS-CoV-2 is still unclear (*5,6*). These concerns are further confounded by potential immunological differences between immunization versus native infection and the evolutionary nature of the virus (*i.e.* virus variants) (*5*).

For these reasons, serological testing or measurement of circulating antibodies is becoming increasingly important in order to determine if an individual has generated antibodies to SARS-CoV-2 as a result of infection and/or vaccination (*4,7*). Serological measurements are typically performed using serum or plasma samples obtained intravenously, however, dried blood spot (DBS) samples offer an alternative means of sample collection with several advantages over traditional phlebotomy. In addition to utilizing a smaller volume of blood (∼50 µL of blood/spot) and less stringent shipping requirements, DBS cards can be successfully collected in the home setting with minimal training (*8–10*).

Since the start of the pandemic, many laboratories and researchers have investigated the use of DBS collection for both qualitative and quantitative detection of SARS-CoV-2 antibodies (*10–12*). These assays have demonstrated high sensitivity and specificity when comparing DBS results to plasma or serum in proof-of-concept studies. However, further investigation including studies based on regulatory guidance is required prior to utilization of DBS samples for at-home self-collection (*13*). In the work presented here, we demonstrate the ability to measure SARS-CoV-2 antibodies from DBS samples using the Roche Elecsys® Anti-SARS-CoV-2 S electrochemiluminescence immunoassay which has received emergency use authorization (EUA) for semi-quantitative measurement of antibodies in venous serum and plasma samples (*14*). The feasibility of using this assay to measure SARS-CoV-2 antibodies has been demonstrated previously (*12*). However, the studies performed herein represent a more rigorous testing of this assay including a simplified extraction process, a reduction in the assay’s reporting limit for DBS samples, and demonstration of sample self-collection. Lastly, serial monitoring of SARS-CoV-2 antibodies was demonstrated using self-collected DBS samples. FDA EUA guidance for serological testing for at-home sample collection was utilized where applicable (*13*).

## 2. Material and Methods

### 2.1. Sample Extraction and Measurement

For extraction of DBS samples, two ¼” diameter round hole punches were taken from regions of the DBS card that were saturated with blood (*i.e.* no white is visible on the punches) and placed into a single 16 x 75 mm polypropylene tube. Punches were then submerged in 150 µl of Roche Universal Diluent (07299001190) using a wooden applicator. Tubes were then placed on a micro plate shaker (VWR, 12620-926) at 240 rpm for one hour at room temperature (20-25°C). Following extraction, remnant solution was squeezed out of the punches which were then discarded. The remaining extract (∼100 µL) was then transferred into a Hitachi microcup (system dead volume of 50 µL) for measurement on a Roche Cobas 8000 e801 immunoassay module.

Measurement of SARS-CoV-2 antibodies was performed using the Roche Elecsys® Anti-SARS-CoV-2 S assay which has received EUA approval for the semi-quantitative measurement of total SARS-CoV-2 antibodies in serum and plasma samples. With the assistance of Roche, the lower numerical reporting limit was reduced from 0.400 U/mL (venous sample limit of quantitation) to 0.000 U/mL. This is necessary as samples obtained using DBS are diluted (∼10-fold) through the extraction process. By reducing the reporting limit, a lower LOQ for DBS extracts could be investigated and reduce the possibility of false negative results for patients with serum antibody results just above the serum clinical cutoff (0.800 U/mL) (*14*). For example, a patient with serum antibody results of 3.00 U/mL would have a DBS result of 0.300 U/mL (assuming 10-fold dilution and complete analyte recovery) which is less than the EUA approved assay LOQ (0.400 U/mL). No other EUA assay parameters were modified.

### 2.2. Clinical Agreement Studies

Paired venous serum and DBS samples were collected from individuals with previous COVID-19 infection (based on EUA approved RT-PCR diagnostic assays, n = 36) as well as from presumed negative individuals (n = 84). Donors previously infected with COVID-19 had nasopharyngeal samples collected and tested (using EUA approved methodologies for detection of SARS-CoV-2) between October and December 2020. All DBS and serum samples for this study were collected in January 2021.

Venous samples were obtained using traditional venipuncture techniques while DBS samples were obtained following sterilization of the fingertip with an alcohol pad and lancing the finger with a high flow contact-activated lancet (BD #366594). After wiping the first drop of blood with a gauze pad, blood was applied to the DBS card (Eastern Business Forms 903™ Dried Blood Spot Card, 10550021) to fill all five spots (∼50 μL/spot). No instructions were provided regarding milking or squeezing of the finger after the fingerstick. Following collection, DBS samples were dried for three hours at room temperature (20-25°C) and then placed into a plastic specimen pouch (without desiccant) for storage until testing. For self-collection of DBS samples, all donors provided samples in a home-like setting using detailed instructions for use to assist with the process described above. To rule out potential active asymptomatic infection for presumed negative donors, a self-collected nasal swab was procured concurrently and analyzed using Labcorp’s EUA approved RT-PCR assay for SARS-COV-2 (*15*).

Participants within this study represented individuals with varying levels of education and self-collection experience. All studies were performed under two Institutional Review Board approved protocols: SQNM-RND-103 (IRB number 520100174, study number 1269845) and SCMM-RND-402 (IRB number 520180046, study number 1270479).

### 2.3. Creation of Contrived Blood DBS Samples

For specific validation studies, contrived blood samples were created due to the difficulty of obtaining DBS samples at specific concentrations and in large volumes. These samples were generated by mixing venous serum (screened for SARS-CoV-2 antibodies) with packed red blood cells to create samples with 40% hematocrit. Red blood cells were obtained intravenously from a seronegative donor with Type O blood using EDTA tubes. After mixing, the contrived blood samples were pipetted onto DBS cards (∼50 μL/spot) which were then allowed to dry for 3 hours at room temperature prior to storage. For many studies (specifically those regarding sample and assay robustness), contrived blood samples were created such that approximately 25% of the samples utilized were negative samples within 5x the DBS assay cutoff, 50% were positive samples within 5x the cutoff, and 25% were greater than 5x the cutoff.

## 3. Results and Discussion

### 3.1. Assessment of DBS Analytical Measurement Range and Imprecision

#### 3.1.1. DBS Limit of Blank (LOB)

In order to assess the detection capability of the assay with DBS extracts, guidance from CLSI EP17-A2 was utilize (*16*). Two separate reagent lots were used to make 96 blank measurements on 6 contrived blood samples over a 4-day period. Two results were not included in the data analysis for each lot as the z-score for each of these results with respect to the remaining results were greater than 4.7 for both reagent lots. Using the mean and standard deviation of the remaining blank results (n = 94) as well as a normal distribution multiplier, the LOB for DBS extracts was determined to be 0.111 U/mL (Supplementary Section 1).

#### 3.1.2. DBS Limit of Detection (LOD)

The limit of detection (LOD) was assessed using 5 contrived blood samples in accordance with CLSI EP05-A3 and CLSI EP17-A2 guidance (Supplementary Section 2) (*16,17*). The samples utilized had mean results (across two reagent lots) that were expected to be close to the clinical cutoff (0.146 to 0.531 U/mL). Pooled standard deviations were calculated for the two different reagent lots using CLSI EP17-A2 guidance (*16*). The first reagent lot produced a pooled standard deviation of 0.0419 U/mL while the second demonstrated a standard deviation of 0.0346 U/mL. Using the larger standard deviation (0.0419 U/mL), a normal distribution multiplier, and the reported LOB (0.111 U/mL) the LOD for DBS extracts was determined to be 0.180 U/mL.

#### 3.1.3. DBS Limit of Quantitation (LOQ)

Measurements to determine the limit of quantitation (LOQ) of DBS extracts utilized 14 contrived blood samples covering an appropriate range of concentrations (0.0528 to 0.648 U/mL) These samples were extracted and measured in triplicate over a five-day period (15 total measurements for each level) using two different reagent lots on a single instrument. For this study, the target CV and bias were set to 25.0% based on FDA guidance for ligand binding assays at the lower limit of quantitation (*18*). Following collection of data, the imprecision profiles were analyzed using the Limit of Quantitation module in EP Evaluator^®^ (Supplementary Section 3). These results indicated an LOQ of 0.0873 U/mL for the first reagent lot while the second reagent lot demonstrated an LOQ of 0.0736 U/mL. In addition, acceptable biases less than ±25.0% were observed for all levels greater than the DBS LOD. As both imprecision and bias results indicate an LOQ less than the observed LOD, the LOQ for DBS extracts is in practice equivalent to the LOD – 0.180 U/mL (*16*).

Investigation of the DBS assay’s LOB, LOD, and LOQ indicates that a lower reporting limit can be achieved for DBS extracts. This may be attributable to the reduction of sample dependent matrix effects as a result of ∼10-fold extraction (dilution) with the approved assay diluent (*19*). As a result of the increased sensitivity, a clinical cutoff value below the EUA approved assay’s LOQ for serum and plasma can be implemented for DBS extracts, enabling assessment of categorical agreement between venous and DBS samples near the serum cutoff (0.8 U/mL).

#### 3.1.4. DBS Linearity

Linearity of the assay with DBS samples was assessed using 11 antibody concentrations made through serum sample admixtures in 10% increments followed by contrivance into DBS (Supplementary Section 4). Initial assessment of linearity indicated acceptable biases (±20.0%) from 0.0667 to 147 U/mL with 147 U/mL being the highest concentration tested. This study was repeated with samples of higher concentration to extend the measurement range to 250 U/mL in order to match the reporting limit of un-diluted samples for the EUA approved assay. For this second study, samples with extracted concentrations that initially measured > 250 U/mL were manually diluted 10-fold with Universal Diluent and re-measured as indicated in the assay’s package insert (*14*). Antibody concentrations for this second study ranged from 0.0974 to 323 U/mL and targets were determined by using a linear fit of all data points.

#### 3.1.5. DBS Clinical Cutoff

The clinical cutoff, used to assign DBS results as negative or positive for antibodies, for DBS samples was created using results from self-collected samples from donors confirmed to be seronegative (n = 77, Supplementary Section 5). The standard deviation of these results (0.0405 U/mL) was multiplied by 3 and added to the mean (0.0625 U/mL) to give a value of 0.184 U/mL. The DBS clinical cutoff for positivity was then set to ≥0.185 U/mL, where all results less than 0.185 are reported as negative.

#### 3.1.6. DBS Analytical Measurement Range

Evaluation of the detection capability of the assay with DBS extracts indicates that sample concentrations can be measured from 0.180 U/mL (LOD/LOQ) to 250 U/mL (upper reporting limit of assay) with a clinical negative/positive antibody cutoff concentration of 0.185 U/mL. This DBS range represents a calculated serum measurement range of 2.6 to 3,570 U/mL. Although dilution of samples is performed for venous serum and plasma samples for the EUA approved in order to extend the reporting limit, dilution of DBS samples is not currently utilized due to the relatively wider concentration range that can be reported for DBS samples (as a result of pre-dilution through extraction).

#### 3.1.7. Imprecision of DBS Samples

Assay imprecision was assessed for DBS extracts using two reagent lots over a 4-day period with 6 contrived blood samples that covered a range of antibody concentrations (0.504 to 156 U/mL). A total of 16 replicates for each sample were measured (Supplementary Section 6). For both reagent lots, the CVs observed for repeatability and within-laboratory imprecision were less than 15.0% which meets FDA specifications for ligand binding assays (Table 1) (*18*). All samples had a total categorical agreement of 100.0%.

**Table 1.** Imprecision of extracted DBS samples (Reagent Lots 53688601 / 54862501).

| Sample | Mean<br>(U/mL) | Repeatability<br>(%) | Within-Laboratory<br>(%) | Total<br>Agreement<br>(%) |
| --- | --- | --- | --- | --- |
| 1 | 0.530 / 0.504 | 7.6 / 6.1 | 10.9 / 9.4 | 100.0 / 100.0 |
| 2 | 0.682 / 0.661 | 8.5 / 7.6 | 11.3 / 10.1 | 100.0 / 100.0 |
| 3 | 5.97 / 5.93 | 7.7 / 7.6 | 14.7 / 12.8 | 100.0 / 100.0 |
| 4 | 15.3 / 15.3 | 6.9 / 6.1 | 8.6 / 8.0 | 100.0 / 100.0 |
| 5 | 61.8 / 61.9 | 12.5 / 13.2 | 12.5 / 13.2 | 100.0 / 100.0 |
| 6 | 156 / 156 | 7.7 / 7.9 | 8.7 / 7.9 | 100.0 / 100.0 |

### 3.2. Clinical Correlation Study

Following collection and measurement of samples, 6 of the 84 presumed negative donors had venous serum, self-collected DBS samples, and professionally collected DBS samples measure positive for SARS-CoV-2 antibodies despite having negative RT-PCR results at the time of serological specimen collection (Supplementary Section 7). The serum results for these donors ranged from 8.81 U/mL to 1170 U/mL where the clinical cutoff for serum samples is 0.800 U/mL. These results suggest previous (asymptomatic) infection or unreported vaccination. To confirm these results, the serum samples for these donors were measured using three additional EUA approved serology assays (Roche Elecsys® Anti-SARS-CoV-2, DiaSorin Liaison SARS-CoV-2 S1/S2 IgG and DiaSorin Liaison SARS-CoV-2 IgM) (*20*). All six donors had serum results measure as positive on at least one additional assay and as such results were excluded from qualitative and quantitative analysis. All remaining results obtained were analyzed qualitatively using the established clinical cutoff for DBS extracts (0.185 U/mL) and quantitatively through correlative analysis.

Comparison of serum samples with DBS obtained through self-collection demonstrated a high degree of agreement (R = 0.96) and Deming slope of 0.069 (n = 108), which is attributed to dilution of the sample through the extraction process as well as precise but incomplete recovery of antibodies (Figure 1A and 1C). Two donors with negative venous serum results had self-collected DBS samples that measured positive for antibodies (results within 3.5x the DBS clinical cutoff, Supplementary Section 7). As a result of these two false positives as well as a false negative donor with unmeasurable antibody results for both serum and DBS, qualitative total categorical agreement of self-collected DBS samples compared to venous serum results was 98.1% (Table 2A). In addition, the negative percent agreement (NPA) and positive percent agreement PPA were found to be 97.4% and 100.0%, respectively. When compared to RT-PCR results, qualitative categorical agreement was 97.2% with an NPA and PPA of 97.3% and 97.1%, respectively (Table 2B). Results below and above the DBS measurement range were interpreted as negative and positive, respectively.

**Figure 1.**
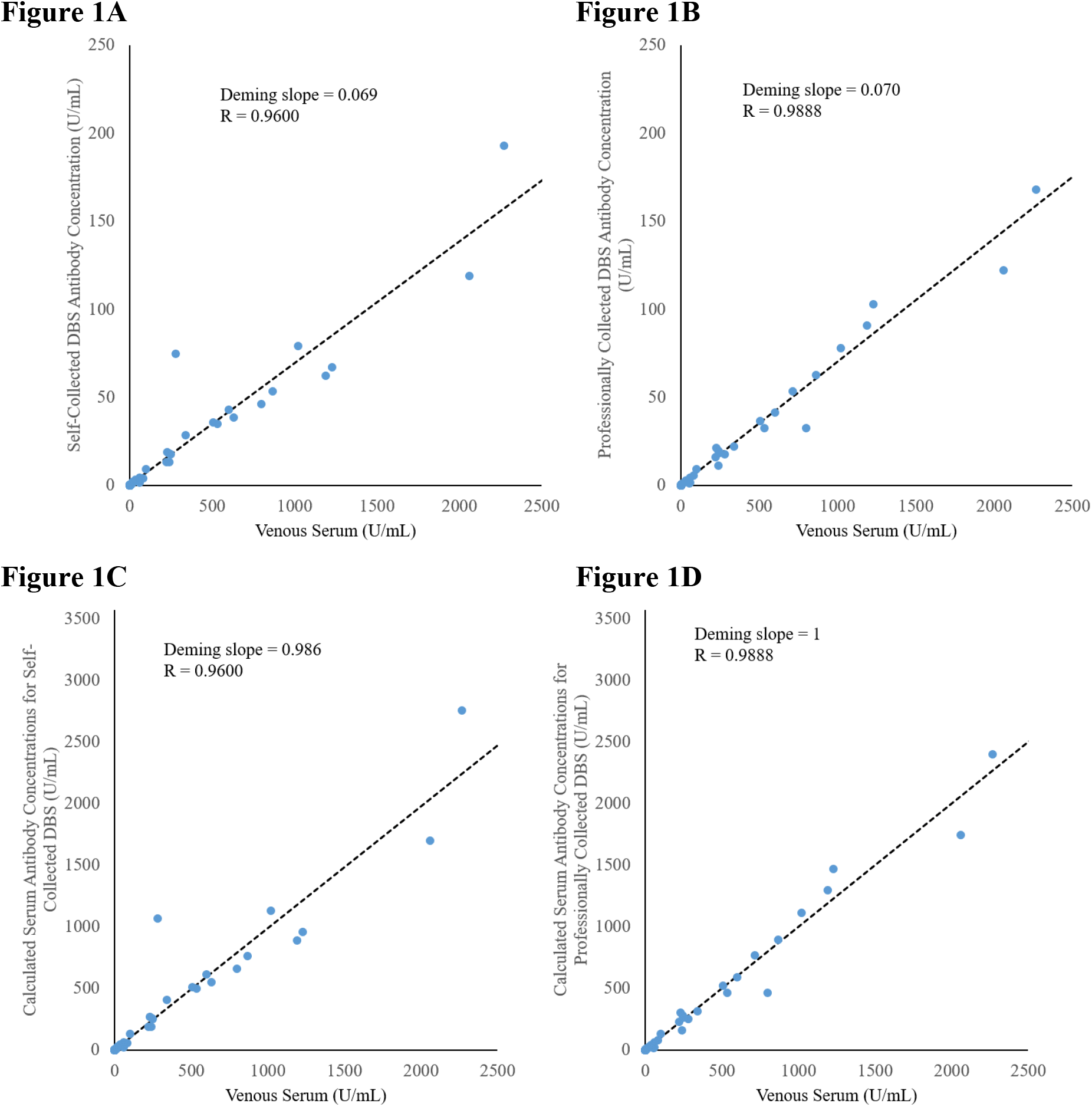
Serum antibody concentrations compared to (A) self-collected DBS sample antibody levels and (B) professionally collected DBS sample antibody levels. Calculated serum concentrations are also shown with respect to DBS concentrations (by dividing DBS results by 0.070) for both (C) self-collected and (D) professionally collected samples.

**Table 2.** Qualitative comparisons of self-collected DBS results to (A) serum and (B) RT-PCR results and professionally collected DBS results to (C) serum and (D) RT-PCR results.

|  |  |  |  |  |  |
| --- | --- | --- | --- | --- | --- |
| **A)** |  | **Serum Results** |  |  |  |
|  |  | - | + |  |  |
| **Self-Collected DBS Results** | - | 74 | 0 | 74 | NPV = 100.0% |
|  | + | 2 | 32 | 34 | PPV = 94.1% |
|  |  | 76 | 32 | 108 | **Total Agreement:** |
|  |  | NPA = 97.4% | PPA = 100.0% |  | **98.1%** |
| **B)** |  | **RT-PCR Results** |  |  |  |
|  |  | - | + |  |  |
| **Self-Collected DBS Results** | - | 73 | 1 | 74 | NPV = 98.6% |
|  | + | 2 | 33 | 35 | PPV = 94.3% |
|  | Total | 75 | 34 | 109 | **Total Agreement:** |
|  |  | NPA = 97.3% | PPA = 97.1% |  | **97.2%** |
| **C)** |  | **Serum Results** |  |  |  |
|  |  | - | + |  |  |
| **Professionally Collected DBS Results** | - | 79 | 0 | 79 | NPV = 100.0% |
|  | + | 0 | 32 | 32 | PPV = 100.0% |
|  |  | 79 | 32 | 111 | **Total Agreement:** |
|  |  | NPA = 100.0% | PPA = 100.0% |  | **100.0%** |
| **D)** |  | **RT-PCR Results** |  |  |  |
|  |  | - | + |  |  |
| **Professionally Collected DBS Results** | - | 78 | 1 | 79 | NPV = 98.7% |
|  | + | 0 | 32 | 32 | PPV = 100.0% |
|  |  | 78 | 33 | 111 | **Total Agreement:** |
|  |  | NPA = 100.0% | PPA = 97.0% |  | **99.1%** |

When quantitatively comparing professionally-collected DBS samples to serum results (n = 106), results were similar to self-collected results with a correlation coefficient (R) of 0.9888 and a Deming slope of 0.070 (Figure 1B and 1D). Total qualitative categorical agreement (n = 111) to venous serum results was 100.0% (Table 2C). When compared to RT-PCR results, qualitative categorical agreement was 99.1% with an NPA and PPA of 100.0% and 97.0%, respectively (Table 2D). These results met FDA guidance at the time of these studies (NPA ≥ 95.0%, PPA ≥ 90.0%) (*13*).

### 3.3. Robustness Studies

In order to evaluate sample stability during the shipping process, simulated shipping studies were performed in accordance with ISTA 7D guidance as recommended by the FDA (*13,21*). Contrived DBS samples were prepared in triplicate and split between three storage conditions: room temperature (20-25°C), a simulated winter and summer shipping excursions (Supplementary Section 8). Following completion of the excursions, samples were measured in a single measurement run where results from samples stored in parallel at room temperature were used as baseline results.

Additional robustness and analytical interference studies were performed to stress different aspects of the DBS sample collection process as well as the influence of several endogenous or exogenous interferences (Supplementary Sections 9 and 10). For acceptance of results, a mean bias of ±20.0% was used as quantitative acceptance following the FDA guidance for ligand binding assays (*18*). For qualitative assessment, a total categorical agreement of 95.0% was utilized based on guidance from the FDA’s Home Specimen Collection Serology Template (*13*).

#### 3.3.1. DBS Shipping Stability

For shipping excursion studies, contrived blood samples were created in triplicate and split into three shipping conditions: baseline ambient (20-25°C), winter and summer excursions (Supplementary Section 8). Results less than 0.180 U/mL (DBS assay LOQ) were included in qualitative analysis but excluded from quantitative analysis. Overall, the sample results from winter and summer excursions both demonstrated total categorical agreement of 97.4% with mean biases of 6.0% and −0.5%, respectively (Table 3). These results indicate that samples are stable from the time of collection in an individual’s home until received in the laboratory.

**Table 3.** Summary of DBS robustness study results.

| **Excursion** | **Hour(s)** | **Mean Bias** | **n\*:** | **Categorical Agreement** | **n:** |
| --- | --- | --- | --- | --- | --- |
| **Winter** | 56 | 6.0% | 58 | 97.4% | 76 |
| **Summer** | 56 | -0.5% | 57 | 97.4% | 76 |
|  | **Hour(s)** | **Mean Bias:** | **n\*:** | **Categorical Agreement:** | **n:** |
| **Alternate Drying Times** | 0 | -32.5% | 12 | 95.0% | 20 |
|  | 1 | 1.3% | 12 | 95.0% | 20 |
|  | 22 | 1.3% | 12 | 100.0% | 20 |
| **Humid Drying** | 1 | -13.3% | 12 | 95.0% | 20 |
|  | 3 | -22.2% | 12 | 95.0% | 20 |
|  | 22 | -44.6% | 12 | 95.0% | 20 |
| **Contamination** |  | **Mean Bias :** | **n\*:** | **Categorical Agreement ;** | **n:** |
| **Alcohol Exposure** |  | -11.0% | 13 | 100.0% | 20 |
| **Pressing Finger to Card** |  | 1.3% | 14 | 95.0% | 20 |

\*Results below LOD were excluded from bias analysis

#### 3.3.2. Stress Testing the Collection Process

For robustness studies, results that were found to be less than the assay’s DBS LOQ were excluded from bias analysis but included in qualitative analysis. Investigation of drying times compared to the recommended drying time of 3 hours (prior to sample storage) was performed. Samples that were immediately stored after contrived blood was added to the card had a categorical agreement of 95.0% and mean bias of −32.5%. Results from samples that were dried for 1 and 22 hours demonstrated categorical agreements of 95.0 and 100.0%, respectively with mean bias of 1.3% for both (Table 3). Effects of drying samples in a humid environment (40°C, 95% RH) were also investigated (*23*). All results observed had a categorical agreement of 95.0%, but the magnitude of the bias increased from −13.3% for 1 hour to −44.6% for 22 hours of humid drying.

Contamination as a result of potential errors in the collection process was investigated (Table 3, Supplementary Section 9). Exposure of DBS spots to alcohol, which may occur following finger sterilization without allowing the fingertip to dry, demonstrated a categorical agreement of 100.0% and mean bias of −11.0%. Contamination of the DBS card by an unsterilized finger prior to collection demonstrated a categorical agreement of 95.0% and mean bias of 1.3%. Although all robustness studies had a total categorical agreement greater than or equal to 95.0%, biases less than ±20.0% were observed in some instances. As these studies are not exhaustive of all possible contaminants, proper instruction materials must be provided to the patient in order to insure the collection of a sample of sufficient quality for measurement.

#### 3.3.3. Analytical Interference Studies

Several studies were performed to assess the effects of different endogenous and exogenous interferents on the measurement of SARS-CoV-2 antibodies from DBS samples (Supplementary Section 10). Results from all interference studies had a categorical agreement greater than or equal to 95.0% to baseline measurements (Table 4). The exogenous interferents tested had mean biases less than ±5.0% while most endogenous interferents tested had a mean bias result less than ±10.0% with the exception of excess protein (17.7%).

**Table 4.** Summary of DBS analytical interference results.

| Analytical Interferents: |  | Mean Bias: | n*: | Categorical Agreement: | n: |
| --- | --- | --- | --- | --- | --- |
| **Endogenous Interferents** | Hemolysis (100%) | 3.6% | 15 | 95.0% | 20 |
|  | Triglycerides (3000 mg/dL) | 4.5% | 18 | 96.7% | 30 |
|  | Total Protein (12 g/dL) | 17.7% | 19 | 100.0% | 30 |
|  | Conjugated Bilirubin (20 mg/dL) | -2.8% | 20 | 96.7% | 30 |
|  | Unconjugated Bilirubin (20 mg/dL) | -8.7% | 20 | 100.0% | 30 |
| **Exogenous Interferents** | Cerilliant Mix 1 | -0.7% | 20 | 96.7% | 30 |
|  | Cerilliant Mix 2 | 2.5% | 20 | 100.0% | 30 |
|  | Biotin (3510 ng/mL) | 4.5% | 21 | 96.7% | 29 |

\*Results below LOD were excluded from bias analysis

### 3.4. Immunization Study

Application of self-collected DBS was performed with serial monitoring of SARS-CoV-2 antibody levels following immunization in 8 donors. Donors periodically performed self-collection of DBS samples pre-vaccination through 19 weeks following initial vaccination. All donors reported receiving the Pfizer-BioNTech COVID-19 vaccine with the second dose occurring exactly 3 weeks following the first dose. As can be seen in Figure 2A below, all donors had negative DBS results for samples collected within the first 9 days following initial immunization. Antibody levels for each donor rose above the DBS cutoff of 0.185 U/mL between days 10 and 16. Antibody levels increased rapidly following the second vaccination dose (Figure 2B), with several donors reaching levels greater than the DBS reporting limit (250 U/mL which is calculated to be 3570 U/mL in serum) (*22,23*). Dramatically lower antibody levels were observed for Donor 7, likely a result of the immunosuppressive medication the donor reported taking for a chronic condition (*24*).

**Figure 2.**
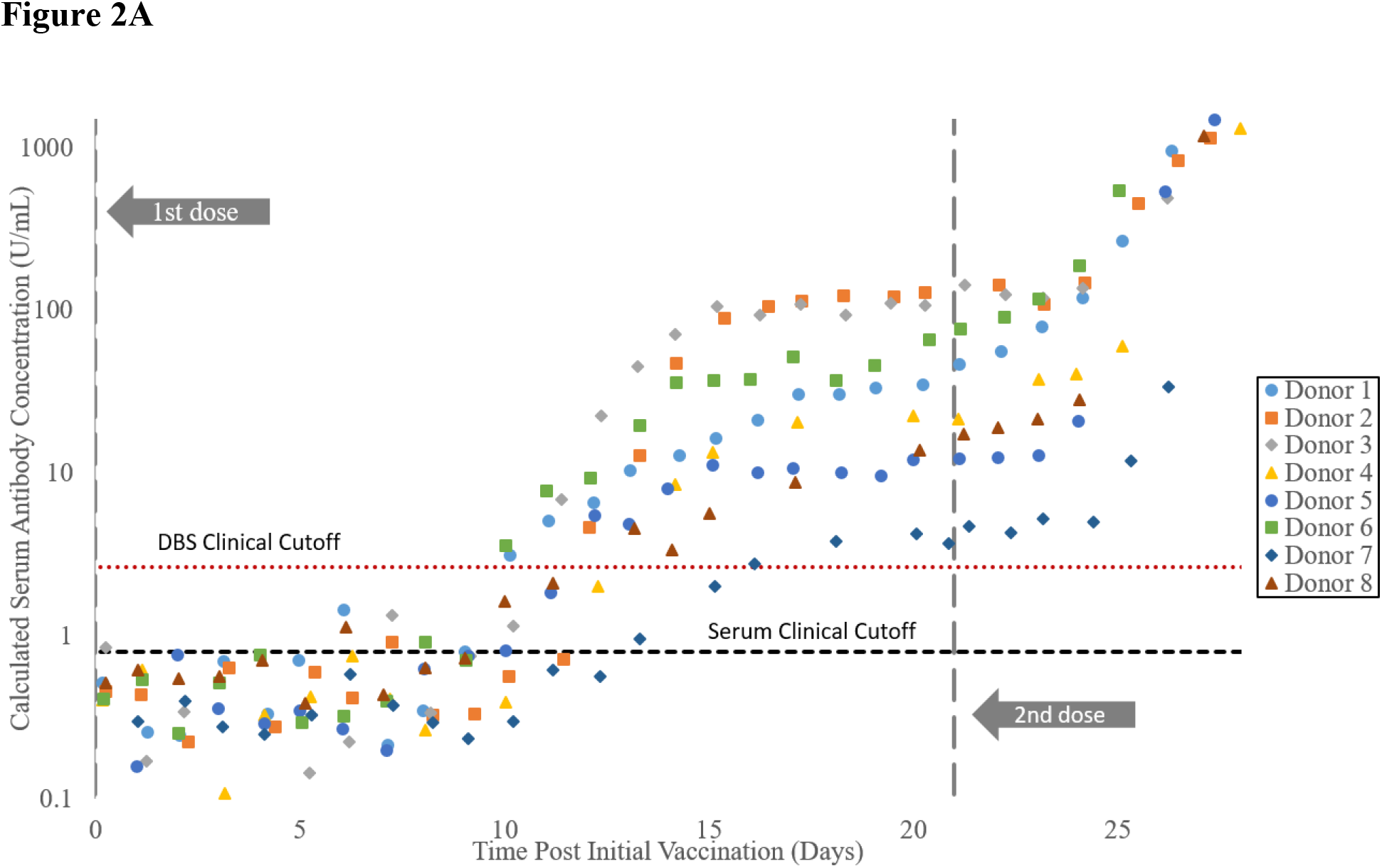
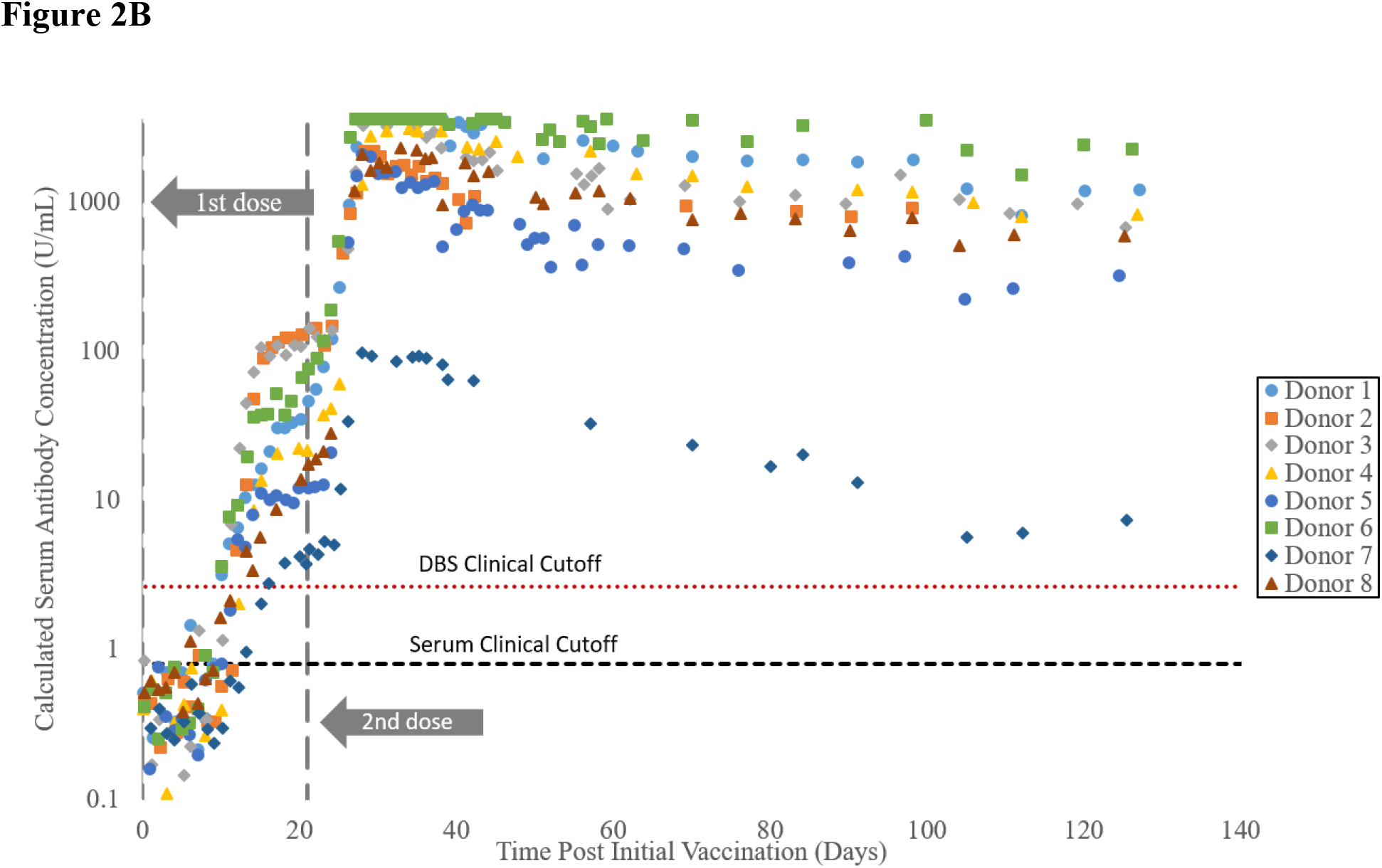
SARS-CoV-2 antibody levels measured from DBS samples following (A) pre-dose (Day 0) through receipt of second vaccine dose (Day 21) to day 28 and (B) through Day 132. Y-axes are displayed logarithmically with the left axis representing calculated serum levels (by dividing DBS results by 0.070).

## 4. Conclusions

The thorough results provided herein demonstrate the robustness of measuring SARS-CoV-2 antibodies using DBS samples. Although DBS samples are diluted through the extraction process (and as a result of incomplete antibody recovery), this approach also has advantages. Using Roche’s Universal Diluent as the extraction buffer, sample-to-sample matrix effects were reduced and a lower (absolute) reporting limit was achieved (LOQ of 0.180 U/mL for DBS samples). In addition, the dilution of the sample allowed a higher relative measurement range to be demonstrated as a DBS samples with a concentration of 250 U/mL DBS sample (upper limit of quantitation for un-diluted venous samples) would be greater than 3,500 U/mL in serum. These results, as well as the high correlation to venous serum results, allowed the assay to be used to demonstrate antibody monitoring over time through at-home DBS self-collections. Ultimately, DBS samples could serve as an important tool for regular antibody monitoring and scheduling of immunization boosters when antibody levels inferring protective immunity is more fully understood in the future.

## Data Availability

All data produced in the present work are contained in the manuscript

## Acknowledgments

The authors appreciate the guidance on serological testing provided by Dr. Alex Katayev, Dr. Laura Gillim, and Dr. Ajay Grover of Laboratory Corporation of America Holdings. In addition, Graham McLennan was instrumental in coordinating donors for clinical studies. The authors would also like to thank Roche Diagnostics particularly Tammy Dean, Stephanie Greeman, Dr. Simon Jochum, Dr. Marcus-Rene Lisy, Dr. Beatus Ofenloch-Haehnle, Reanna Toney, and Daniel Whisenhunt for their assistance and guidance during the setup and testing of this assay with DBS samples.

This work was funded by Laboratory Corporation of America Holdings.

## Author Bio

Peyton Miesse works for Labcorp in the Research and Development Department as a Research Associate II. She received her Master of Science in Biomedical Engineering with a focus in wearable devices in 2020 from Old Dominion University. Her primary research interests include micro-sampling devices intended for self-collection.

## Supplementary Information

### 1. Limit of Blank (LOB)

Six pre-pandemic serum samples were utilized to assess the assay’s limit of blank (LOB) following creation into contrived blood samples for spotting and extraction from DBS cards. This produced 16 total measurements for each sample for a total of 96 “blank” measurements for each lot. A single instrument was used for this study. In accordance with CLSI EP17-A2 guidance, the mean (*M_blank_*) and standard deviation (*SD_blank_*) of all samples was determined across two reagent lots (*1*). All results from sample 4 on Day 2, Run 1 were excluded as the z-score of these results was greater than 4.7 with respect to other results measured using the same lot. In addition to the mean and standard deviation, a multiplier (*c*_*p*_) which represents the 95^th^ percentile of normal distribution was utilized. This multiplier was corrected for the use of the biased observed standard deviation estimate using equation 1 below where B represents the total number of blank results in the dataset (B = 94) and K represents the number of blank samples (K = 6).

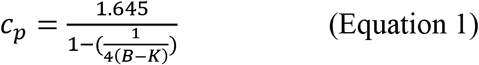

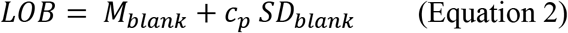

The mean and standard deviation observed for reagent lot 53688601 were 0.0617 and 0.0296 U/mL, respectively (Table 1). With a multiplier value of 1.6497, the calculated LOB for reagent lot 53688601 was 0.111 U/mL. For reagent lot 54862501, the mean and standard deviation was determined to be 0.0417 and 0.0298 U/mL, respectively, giving a LOB of 0.0908 U/mL (Table 2). As the calculated LOB for reagent lot 53688601 is greater, it represents the LOB listed in the manuscript.

**Table 1.**
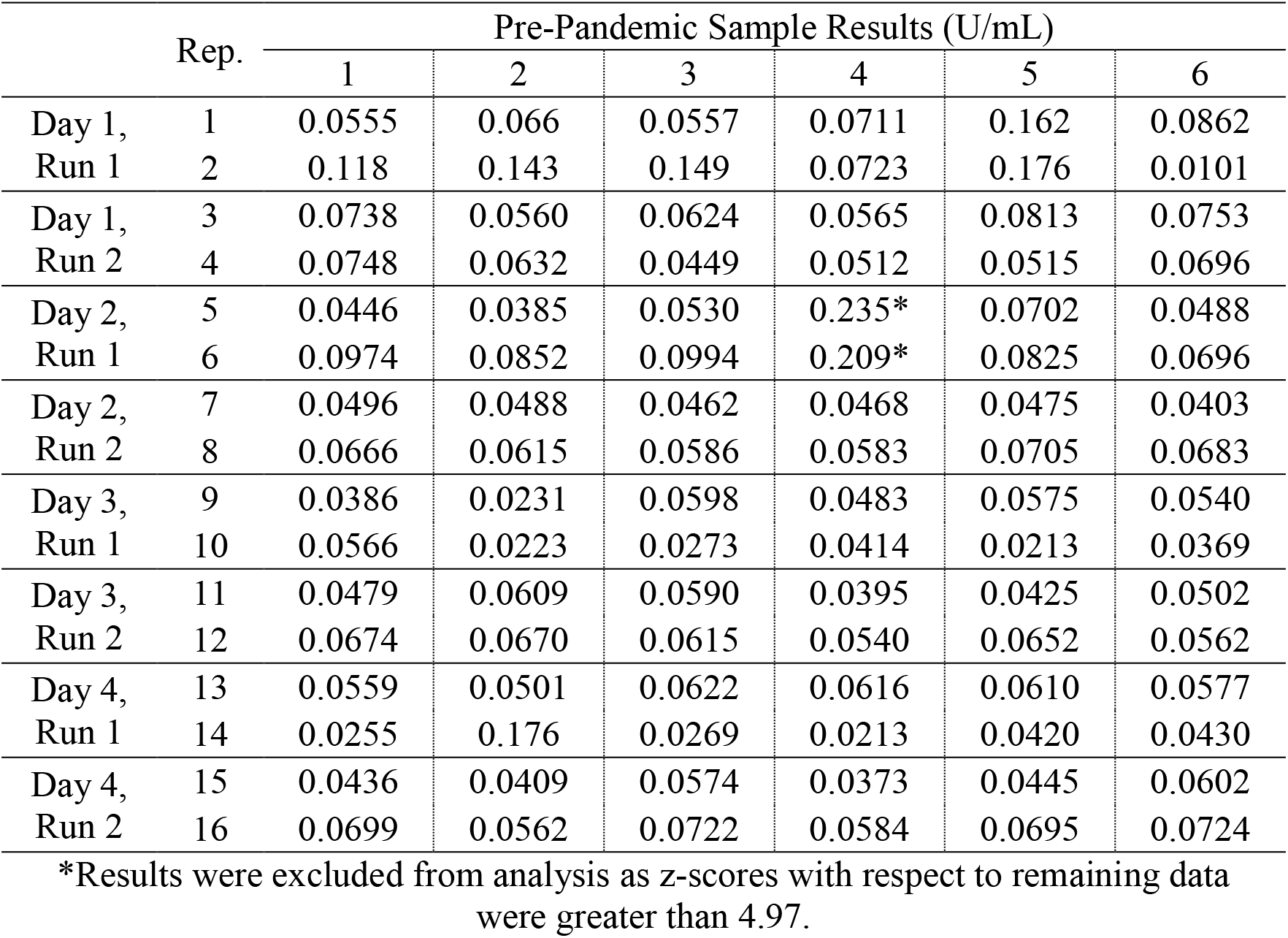
Pre-pandemic DBS limit of blank samples measured with reagent lot 53688601.

**Table 2.**
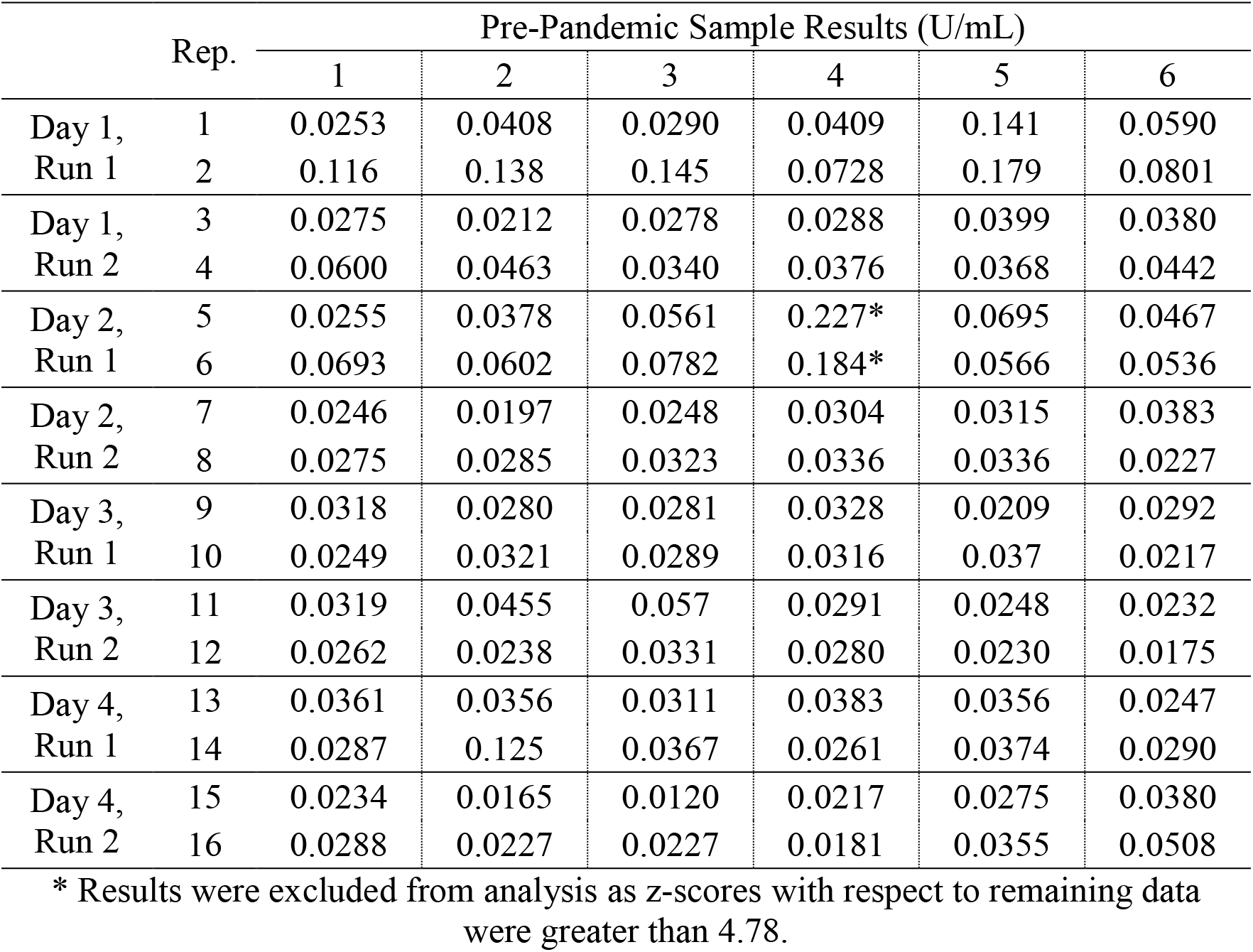
Pre-pandemic DBS limit of blank samples measured with reagent lot 54862501.

### 2. DBS Limit of Detection (LOD)

Five (5) contrived DBS samples were created to have concentrations that were expected to cover the clinical cutoff range (0.126 to 0.415 U/mL), which allows assessment of the assay’s LOB. DBS extracted and measured in duplicate for each run with 2 runs being performed per day over 4 days giving 16 total measurements for each sample. A single instrument was used for this study. In accordance with CLSI EP05-A3 and EP17-A2 guidance, the within laboratory standard deviation of all samples was determined across two reagent lots (*1*). The pooled standard deviation (*SD*_*L*_) was then determined using (*2*):

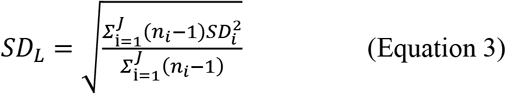

where *SD*_*i*_ represents the SD of all results for the *i*th low level sample, *n*_*i*_ is the number of results for the *i*th low level sample (*n*_*i*_ = 16), and *J* is the number of low level samples (*J* = 5). Using equation 3, the pooled standard deviation for reagent lot 53688601 was found to be 0.0419 U/mL (Table 3) and 0.0346 U/mL for reagent lot 54862501 (Table 4). The larger of these values, 0.0419 U/mL was used with LOB results and a multiplier to determine the assay’s LOD:

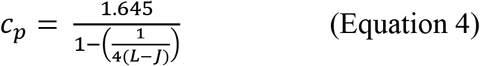

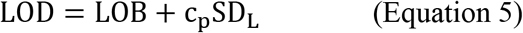

where *L* is the total number of all low level sample (*L* = 80). The LOD was calculated with a multiplier of 1.6505, and the reported LOB (0.111 U/mL) to give a LOD of 0.180 U/mL.

**Table 3.** DBS Limit of detection results (reagent lot 53688601)

| Day # | Rep. | Low Concentration DBS Results (U/mL) |  |  |  |  |
| --- | --- | --- | --- | --- | --- | --- |
|  |  | 1 | 2 | 3 | 4 | 5 |
| Day 1, Run 1 | 1 | 0.170 | 0.182 | 0.279 | 0.325 | 0.382 |
|  | 2 | 0.158 | 0.171 | 0.294 | 0.341 | 0.329 |
| Day 1, Run 2 | 3 | 0.173 | 0.256 | 0.324 | 0.415 | 0.411 |
|  | 4 | 0.146 | 0.188 | 0.332 | 0.349 | 0.531 |
| Day 2, Run 1 | 5 | 0.244 | 0.149 | 0.352 | 0.322 | 0.451 |
|  | 6 | 0.248 | 0.215 | 0.275 | 0.408 | 0.391 |
| Day 2, Run 2 | 7 | 0.204 | 0.202 | 0.303 | 0.365 | 0.390 |
|  | 8 | 0.184 | 0.159 | 0.325 | 0.325 | 0.400 |
| Day 3, Run 1 | 9 | 0.154 | 0.197 | 0.297 | 0.346 | 0.373 |
|  | 10 | 0.169 | 0.153 | 0.300 | 0.338 | 0.370 |
| Day 3, Run 2 | 11 | 0.164 | 0.195 | 0.264 | 0.322 | 0.421 |
|  | 12 | 0.175 | 0.206 | 0.321 | 0.311 | 0.409 |
| Day 4, Run 1 | 13 | 0.170 | 0.181 | 0.265 | 0.320 | 0.354 |
|  | 14 | 0.149 | 0.22 | 0.264 | 0.349 | 0.311 |
| Day 4, Run 2 | 15 | 0.154 | 0.201 | 0.270 | 0.365 | 0.322 |
|  | 16 | 0.160 | 0.168 | 0.232 | 0.324 | 0.333 |
| Total Mean |  | 0.176 | 0.190 | 0.294 | 0.345 | 0.386 |
| Total SD |  | 0.0307 | 0.0277 | 0.0318 | 0.0304 | 0.0548 |
| Total CV |  | 17.4% | 14.5% | 10.8% | 8.8% | 14.2% |
| Repeatability<br>(Error) | SS | 0.00104 | 0.00739 | 0.00507 | 0.00737 | 0.0115 |
|  | DF | 8 | 8 | 8 | 8 | 8 |
|  | MS | 0.000130 | 0.000924 | 0.000634 | 0.000921 | 0.00143 |
|  | SD | 0.0114 | 0.0304 | 0.0252 | 0.0303 | 0.0379 |
| Run (Day) | SS | 0.00279 | 0.00298 | 0.00194 | 0.00355 | 0.0159 |
|  | DF | 4 | 4 | 4 | 4 | 4 |
|  | MS | 0.000699 | 0.000745 | 0.000485 | 0.000888 | 0.00398 |
|  | SD | 0.0264 | 0.0273 | 0.0220 | 0.0298 | 0.0631 |
| Day | SS | 0.0103 | 0.000693 | 0.00752 | 0.00214 | 0.0177 |
|  | DF | 2 | 2 | 2 | 2 | 2 |
|  | MS | 0.00513 | 0.000347 | 0.00376 | 0.00107 | 0.00883 |
|  | SD | 0.0716 | 0.0186 | 0.0613 | 0.0327 | 0.0940 |
| Variance | Error | 0.000130 | 0.000924 | 0.000634 | 0.000921 | 0.00143 |
|  | Run | 0.000284 | 0 | 0 | 0 | 0.00127 |
|  | Day | 1.11E-03 | 0 | 8.19E-04 | 4.51E-05 | 1.21E-03 |
| Within-Laboratory<br>Precision | SD | 0.0390 | 0.0304 | 0.0381 | 0.0311 | 0.0626 |

**Table 4.**
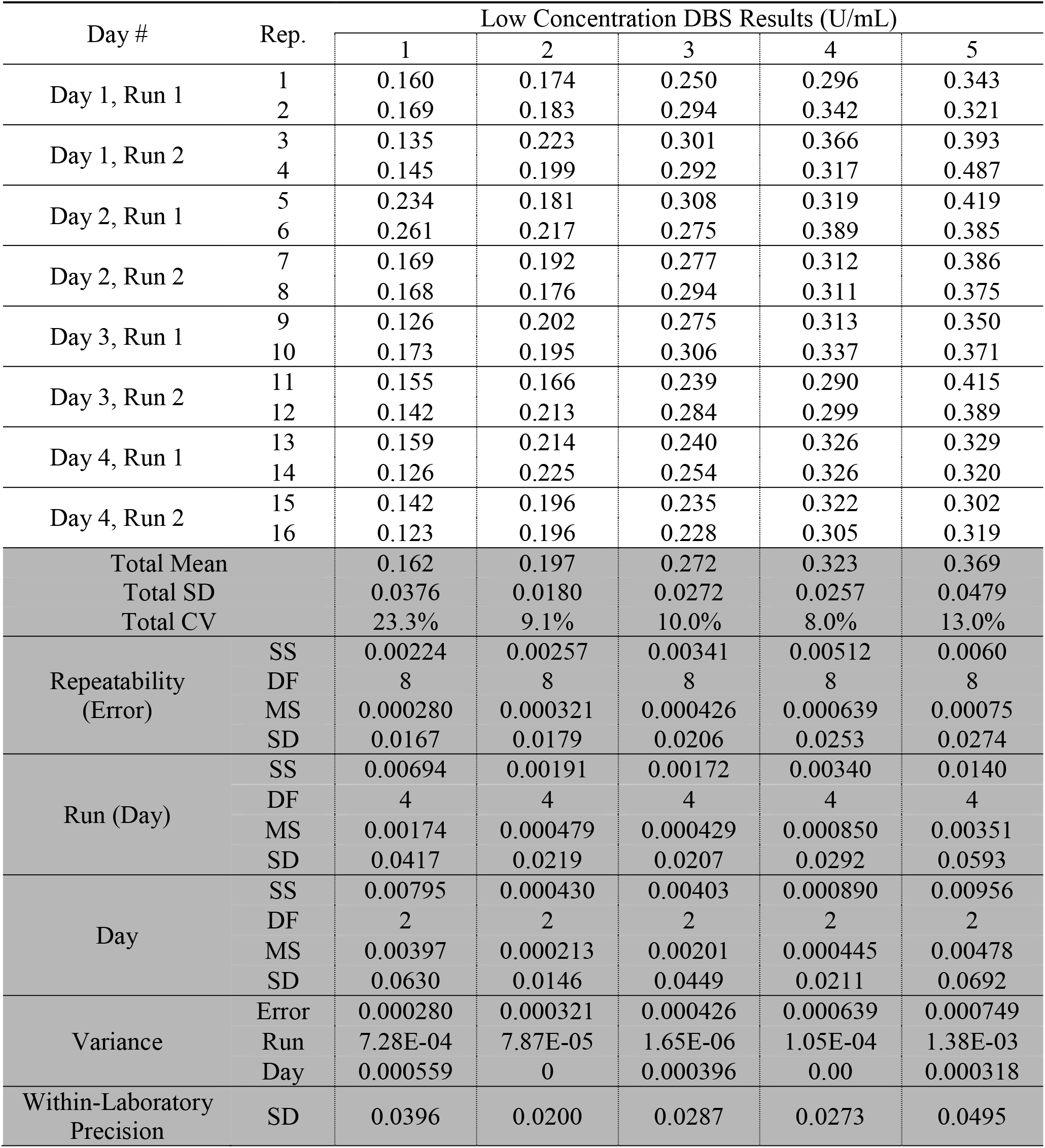
DBS Limit of detection results (reagent tot 54862501)

| Day # | Rep. | Low Concentration DBS Results (U/mL) |  |  |  |  |
| --- | --- | --- | --- | --- | --- | --- |
|  |  | 1 | 2 | 3 | 4 | 5 |
| Day 1, Run 1 | 1 | 0.160 | 0.174 | 0.250 | 0.296 | 0.343 |
|  | 2 | 0.169 | 0.183 | 0.294 | 0.342 | 0.321 |
| Day 1, Run 2 | 3 | 0.135 | 0.223 | 0.301 | 0.366 | 0.393 |
|  | 4 | 0.145 | 0.199 | 0.292 | 0.317 | 0.487 |
| Day 2, Run 1 | 5 | 0.234 | 0.181 | 0.308 | 0.319 | 0.419 |
|  | 6 | 0.261 | 0.217 | 0.275 | 0.389 | 0.385 |
| Day 2, Run 2 | 7 | 0.169 | 0.192 | 0.277 | 0.312 | 0.386 |
|  | 8 | 0.168 | 0.176 | 0.294 | 0.311 | 0.375 |
| Day 3, Run 1 | 9 | 0.126 | 0.202 | 0.275 | 0.313 | 0.350 |
|  | 10 | 0.173 | 0.195 | 0.306 | 0.337 | 0.371 |
| Day 3, Run 2 | 11 | 0.155 | 0.166 | 0.239 | 0.290 | 0.415 |
|  | 12 | 0.142 | 0.213 | 0.284 | 0.299 | 0.389 |
| Day 4, Run 1 | 13 | 0.159 | 0.214 | 0.240 | 0.326 | 0.329 |
|  | 14 | 0.126 | 0.225 | 0.254 | 0.326 | 0.320 |
| Day 4, Run 2 | 15 | 0.142 | 0.196 | 0.235 | 0.322 | 0.302 |
|  | 16 | 0.123 | 0.196 | 0.228 | 0.305 | 0.319 |
| Total Mean |  | 0.162 | 0.197 | 0.272 | 0.323 | 0.369 |
| Total SD |  | 0.0376 | 0.0180 | 0.0272 | 0.0257 | 0.0479 |
| Total CV |  | 23.3% | 9.1% | 10.0% | 8.0% | 13.0% |
| Repeatability<br>(Error) | SS | 0.00224 | 0.00257 | 0.00341 | 0.00512 | 0.0060 |
|  | DF | 8 | 8 | 8 | 8 | 8 |
|  | MS | 0.000280 | 0.000321 | 0.000426 | 0.000639 | 0.00075 |
|  | SD | 0.0167 | 0.0179 | 0.0206 | 0.0253 | 0.0274 |
| Run (Day) | SS | 0.00694 | 0.00191 | 0.00172 | 0.00340 | 0.0140 |
|  | DF | 4 | 4 | 4 | 4 | 4 |
|  | MS | 0.00174 | 0.000479 | 0.000429 | 0.000850 | 0.00351 |
|  | SD | 0.0417 | 0.0219 | 0.0207 | 0.0292 | 0.0593 |
| Day | SS | 0.00795 | 0.000430 | 0.00403 | 0.000890 | 0.00956 |
|  | DF | 2 | 2 | 2 | 2 | 2 |
|  | MS | 0.00397 | 0.000213 | 0.00201 | 0.000445 | 0.00478 |
|  | SD | 0.0630 | 0.0146 | 0.0449 | 0.0211 | 0.0692 |
| Variance | Error | 0.000280 | 0.000321 | 0.000426 | 0.000639 | 0.000749 |
|  | Run | 7.28E-04 | 7.87E-05 | 1.65E-06 | 1.05E-04 | 1.38E-03 |
|  | Day | 0.000559 | 0 | 0.000396 | 0.00 | 0.000318 |
| Within-Laboratory<br>Precision | SD | 0.0396 | 0.0200 | 0.0287 | 0.0273 | 0.0495 |

### 3. DBS Limit of Quantitation (LOQ)

Limit of quantitation (LOQ) studies utilized 14 levels of contrived blood samples that ranged from less than the LOB and ranges that were expected to cover the clinical cutoff (0.0528 to 0.648 U/mL). DBS samples were extracted and measured in triplicate over a five-day period (total of 15 measurements per level). A single instrument was used for this study. Data was analyzed using the Limit of Quantitation module in EP Evaluator® where the imprecision profile is fit to the equation CV = A + B x (1 / mean of measured values). The LOQ is the point where the upper 95% confidence interval of this fit is equal to the target CV. For this application, the target CV was set to 25% based on the FDA guidance for ligand binding assays at the lower limit of quantitation (*3*). Based on imprecision results, reagent lot 51595100 had a LOQ of 0.0873 U/mL while reagent lot 53688600 had a LOQ value of 0.0736 U/mL (Figures 1 and 2). Both of these results are less than the assay’s LOD.

In addition, LOQ results were evaluated in terms of bias where acceptable biases were also set to 25% based on FDA guidance. Bias results (Tables 5 and 6) indicated acceptable bias for all levels greater than the assay’s LOD. As both imprecision and bias results indicated an LOQ less than the assay’s LOD, the reported LOQ for the assay is in practice equivalent to the LOD (0.180 U/mL).

**Figure 1.**
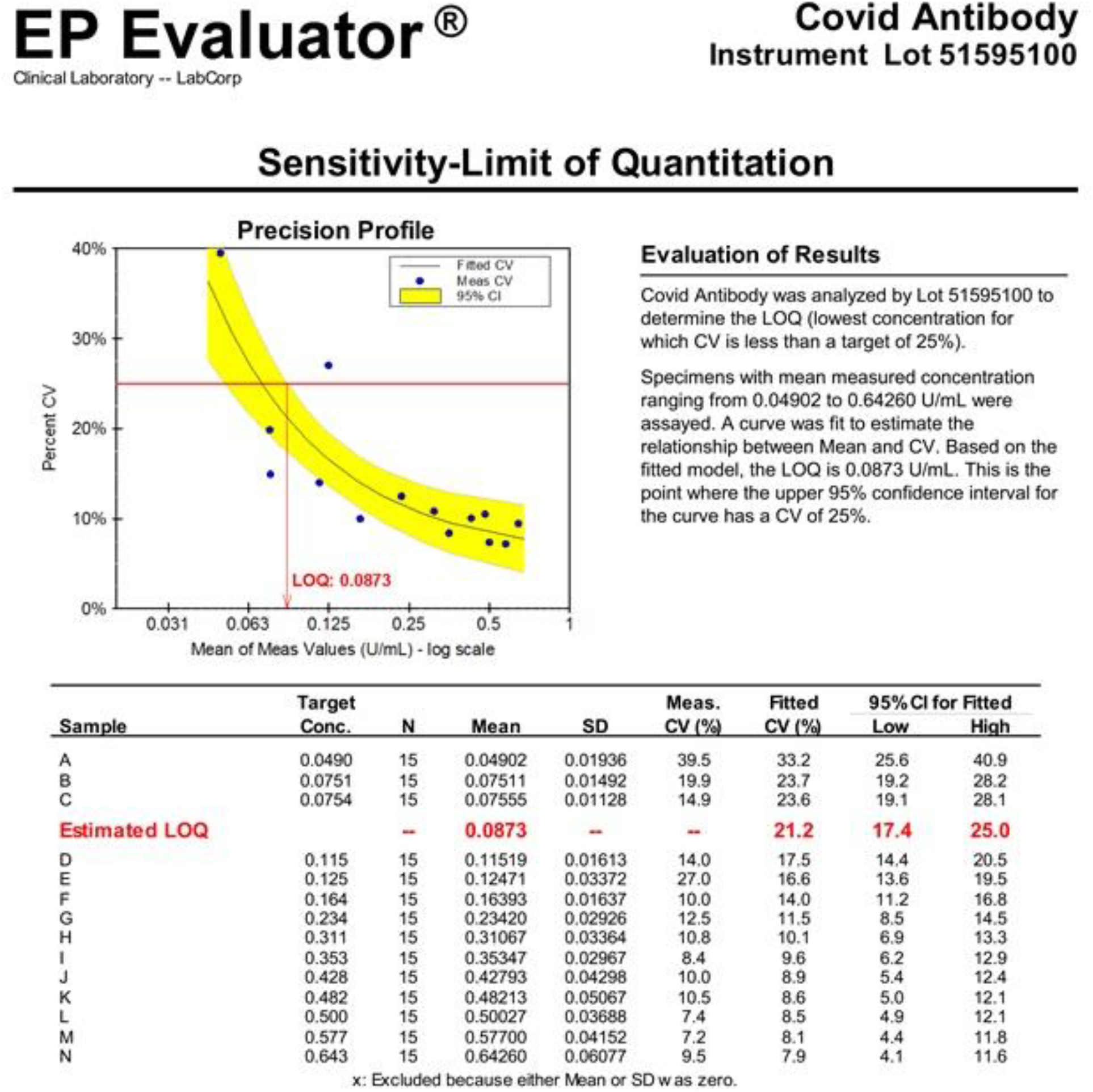
DBS Limit of quantitation imprecision results (reagent lot 51595100)

**Table 5.** DBS Limit of quantitation bias results (reagent lot 51595100)

|  |  | LOQ Sample Results (U/mL) |  |  |  |  |  |  |  |  |  |  |  |  |  |  |  |  |
| --- | --- | --- | --- | --- | --- | --- | --- | --- | --- | --- | --- | --- | --- | --- | --- | --- | --- | --- |
| Level |  | Day 1 |  |  | Day 2 |  |  | Day 3 |  |  | Day 4 |  |  | Day 5 |  |  | Linearity |  |
|  |  | 1 | 2 | 3 | 4 | 5 | 6 | 7 | 8 | 9 | 10 | 11 | 12 | 13 | 14 | 15 | Target | Bias |
| A | 0.0% | 0.0579 | 0.0274 | 0.0498 | 0.0575 | 0.032 | 0.055 | 0.0767 | 0.0725 | 0.0876 | 0.0404 | 0.0440 | 0.0336 | 0.0287 | 0.0505 | 0.0217 | 0.0682 | NA |
| B | 2.5% | 0.0580 | 0.0681 | 0.0951 | 0.0669 | 0.0982 | 0.0649 | 0.0666 | 0.0859 | 0.0976 | 0.0926 | 0.0707 | 0.0796 | 0.0608 | 0.0581 | 0.0635 | 0.0823 | NA |
| C | 5.0% | 0.0729 | 0.0813 | 0.0584 | 0.087 | 0.0904 | 0.0831 | 0.0874 | 0.0873 | 0.0714 | 0.0668 | 0.0869 | 0.0647 | 0.0656 | 0.0575 | 0.0705 | 0.0964 | NA |
| D | 7.5% | 0.107 | 0.0908 | 0.131 | 0.127 | 0.140 | 0.0841 | 0.131 | 0.117 | 0.134 | 0.110 | 0.112 | 0.101 | 0.104 | 0.119 | 0.120 | 0.111 | NA |
| E | 10% | 0.112 | 0.113 | 0.113 | 0.126 | 0.0987 | 0.118 | 0.139 | 0.128 | 0.110 | 0.104 | 0.120 | 0.125 | 0.241 | 0.111 | 0.112 | 0.125 | NA |
| F | 20% | 0.167 | 0.161 | 0.154 | 0.161 | 0.169 | 0.194 | 0.185 | 0.171 | 0.180 | 0.150 | 0.163 | 0.125 | 0.160 | 0.169 | 0.150 | 0.181 | -9.6% |
| G | 30% | 0.205 | 0.263 | 0.194 | 0.234 | 0.267 | 0.244 | 0.280 | 0.271 | 0.247 | 0.238 | 0.182 | 0.230 | 0.232 | 0.224 | 0.202 | 0.238 | -1.5% |
| H | 40% | 0.289 | 0.470 | 0.303 | 0.369 | 0.299 | 0.316 | 0.319 | 0.313 | 0.400 | 0.291 | 0.293 | 0.310 | 0.316 | 0.275 | 0.294 | 0.294 | 10.0% |
| I | 50% | 0.360 | 0.340 | 0.382 | 0.333 | 0.385 | 0.388 | 0.333 | 0.401 | 0.332 | 0.341 | 0.382 | 0.336 | 0.371 | 0.308 | 0.310 | 0.351 | 0.7% |
| J | 60% | 0.447 | 0.468 | 0.625 | 0.472 | 0.452 | 0.427 | 0.489 | 0.425 | 0.432 | 0.320 | 0.400 | 0.421 | 0.438 | 0.430 | 0.358 | 0.407 | 8.1% |
| K | 70% | 0.481 | 0.515 | 0.526 | 0.505 | 0.568 | 0.530 | 0.479 | 0.513 | 0.488 | 0.514 | 0.405 | 0.463 | 0.438 | 0.396 | 0.411 | 0.464 | 3.9% |
| L | 80% | 0.443 | 0.514 | 0.587 | 0.497 | 0.492 | 0.509 | 0.463 | 0.539 | 0.533 | 0.477 | 0.467 | 0.520 | 0.477 | 0.468 | 0.518 | 0.520 | -3.9% |
| M | 90% | 0.541 | 0.559 | 0.579 | 0.601 | 0.607 | 0.637 | 0.599 | 0.554 | 0.530 | 0.617 | 0.58 | 0.654 | 0.527 | 0.553 | 0.517 | 0.577 | 0.0% |
| N | 100% | 0.735 | 0.666 | 0.622 | 0.643 | 0.701 | 0.634 | 0.628 | 0.636 | 0.700 | 0.616 | 0.718 | 0.653 | 0.547 | 0.637 | 0.503 | 0.634 | 1.4% |

Levels E and M used to set Target Values

**Figure 2.**
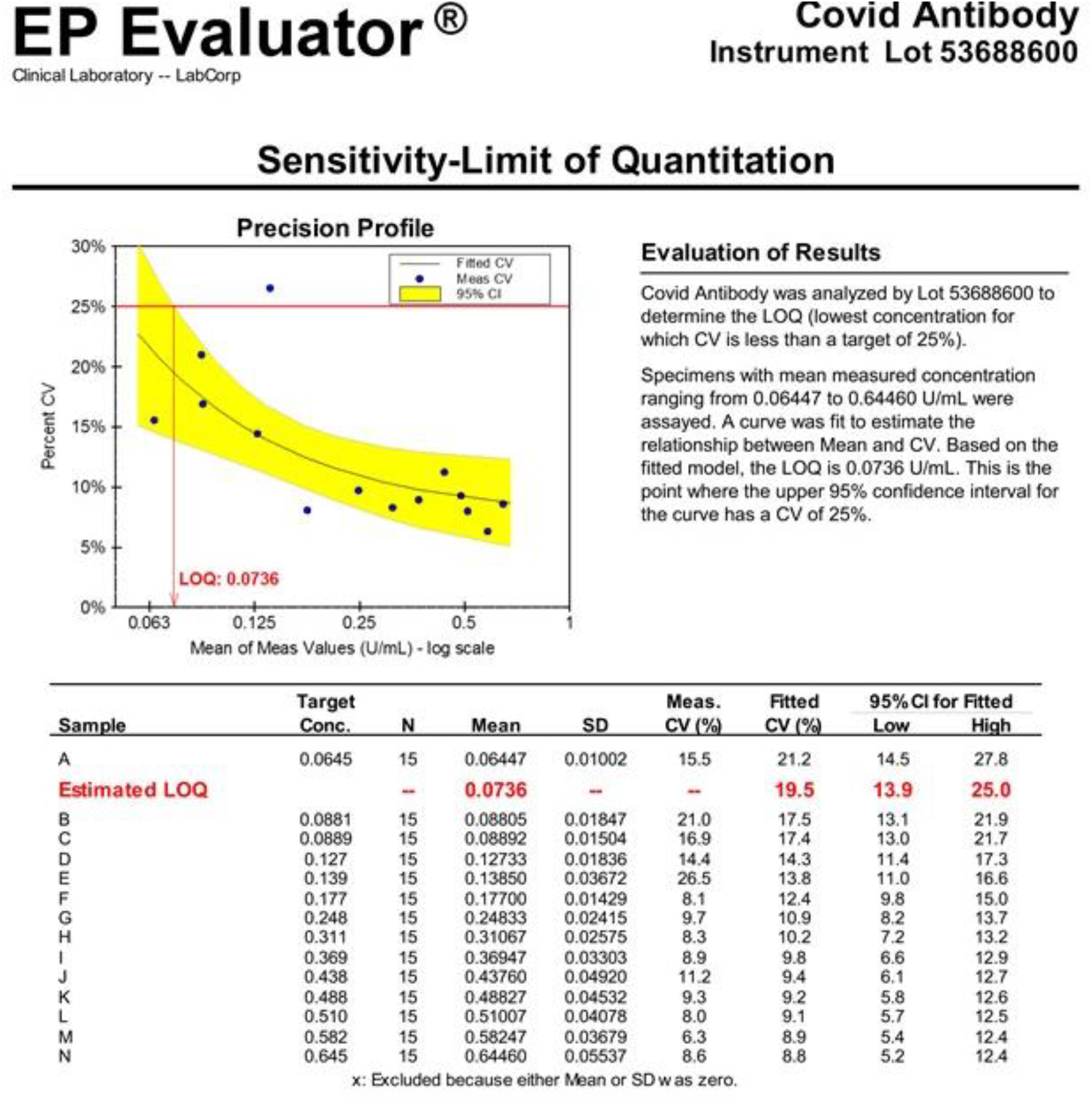
DBS Limit of quantitation imprecision results (reagent lot 53688600)

**Table 6.**
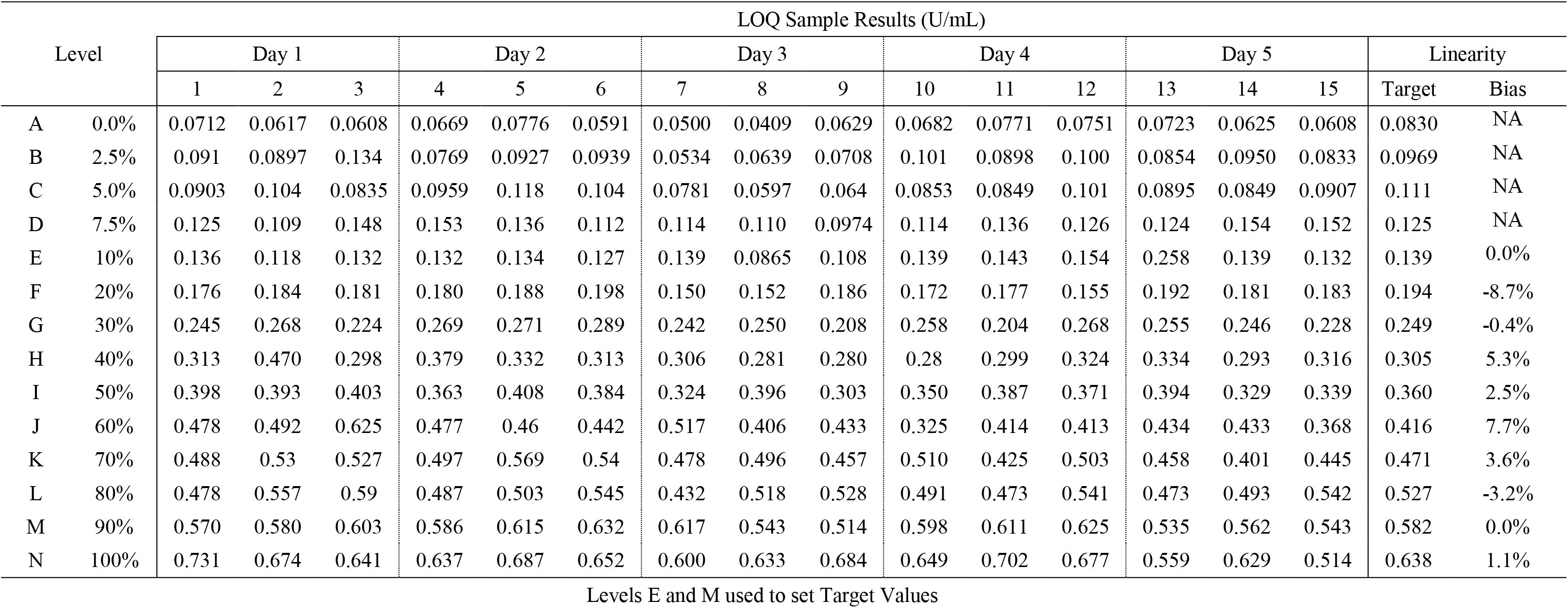
DBS Limit of quantitation bias results (reagent lot 53688600)

| Level |  | LOQ Sample Results (U/mL) |  |  |  |  |  |  |  |  |  |  |  |  |  |  |  |  |
| --- | --- | --- | --- | --- | --- | --- | --- | --- | --- | --- | --- | --- | --- | --- | --- | --- | --- | --- |
|  |  | Day 1 |  |  | Day 2 |  |  | Day 3 |  |  | Day 4 |  |  | Day 5 |  |  | Linearity |  |
|  |  | 1 | 2 | 3 | 4 | 5 | 6 | 7 | 8 | 9 | 10 | 11 | 12 | 13 | 14 | 15 | Target | Bias |
| A | 0.0% | 0.0712 | 0.0617 | 0.0608 | 0.0669 | 0.0776 | 0.0591 | 0.0500 | 0.0409 | 0.0629 | 0.0682 | 0.0771 | 0.0751 | 0.0723 | 0.0625 | 0.0608 | 0.0830 | NA |
| B | 2.5% | 0.091 | 0.0897 | 0.134 | 0.0769 | 0.0927 | 0.0939 | 0.0534 | 0.0639 | 0.0708 | 0.101 | 0.0898 | 0.100 | 0.0854 | 0.0950 | 0.0833 | 0.0969 | NA |
| C | 5.0% | 0.0903 | 0.104 | 0.0835 | 0.0959 | 0.118 | 0.104 | 0.0781 | 0.0597 | 0.064 | 0.0853 | 0.0849 | 0.101 | 0.0895 | 0.0849 | 0.0907 | 0.111 | NA |
| D | 7.5% | 0.125 | 0.109 | 0.148 | 0.153 | 0.136 | 0.112 | 0.114 | 0.110 | 0.0974 | 0.114 | 0.136 | 0.126 | 0.124 | 0.154 | 0.152 | 0.125 | NA |
| E | 10% | 0.136 | 0.118 | 0.132 | 0.132 | 0.134 | 0.127 | 0.139 | 0.0865 | 0.108 | 0.139 | 0.143 | 0.154 | 0.258 | 0.139 | 0.132 | 0.139 | 0.0% |
| F | 20% | 0.176 | 0.184 | 0.181 | 0.180 | 0.188 | 0.198 | 0.150 | 0.152 | 0.186 | 0.172 | 0.177 | 0.155 | 0.192 | 0.181 | 0.183 | 0.194 | -8.7% |
| G | 30% | 0.245 | 0.268 | 0.224 | 0.269 | 0.271 | 0.289 | 0.242 | 0.250 | 0.208 | 0.258 | 0.204 | 0.268 | 0.255 | 0.246 | 0.228 | 0.249 | -0.4% |
| H | 40% | 0.313 | 0.470 | 0.298 | 0.379 | 0.332 | 0.313 | 0.306 | 0.281 | 0.280 | 0.28 | 0.299 | 0.324 | 0.334 | 0.293 | 0.316 | 0.305 | 5.3% |
| I | 50% | 0.398 | 0.393 | 0.403 | 0.363 | 0.408 | 0.384 | 0.324 | 0.396 | 0.303 | 0.350 | 0.387 | 0.371 | 0.394 | 0.329 | 0.339 | 0.360 | 2.5% |
| J | 60% | 0.478 | 0.492 | 0.625 | 0.477 | 0.46 | 0.442 | 0.517 | 0.406 | 0.433 | 0.325 | 0.414 | 0.413 | 0.434 | 0.433 | 0.368 | 0.416 | 7.7% |
| K | 70% | 0.488 | 0.53 | 0.527 | 0.497 | 0.569 | 0.54 | 0.478 | 0.496 | 0.457 | 0.510 | 0.425 | 0.503 | 0.458 | 0.401 | 0.445 | 0.471 | 3.6% |
| L | 80% | 0.478 | 0.557 | 0.59 | 0.487 | 0.503 | 0.545 | 0.432 | 0.518 | 0.528 | 0.491 | 0.473 | 0.541 | 0.473 | 0.493 | 0.542 | 0.527 | -3.2% |
| M | 90% | 0.570 | 0.580 | 0.603 | 0.586 | 0.615 | 0.632 | 0.617 | 0.543 | 0.514 | 0.598 | 0.611 | 0.625 | 0.535 | 0.562 | 0.543 | 0.582 | 0.0% |
| N | 100% | 0.731 | 0.674 | 0.641 | 0.637 | 0.687 | 0.652 | 0.600 | 0.633 | 0.684 | 0.649 | 0.702 | 0.677 | 0.559 | 0.629 | 0.514 | 0.638 | 1.1% |

Levels E and M used to set Target Values

### 4. DBS Linearity

Linearity was performed using positive and negative serum pools that were admixed in 10% increments to create 11 different serum concentrations of SARS-CoV-2 antibodies. Contrived blood samples were created using these different admixtures and applied to DBS cards. Following extraction and measurement, the initial linearity results indicated a linearity from 0.0677 to 147 U/mL for DBS samples (Table 7). Additional linearity results indicated acceptable biases (≤ 20.0%) for all concentrations measured between 0.0974 and 323 U/mL for DBS samples (Table 8). DBS samples that initially measured greater than 250 U/mL were manually diluted 10-fold with Universal Diluent and re-measured.

**Table 7.** Initial DBS linearity results.

| Linearity Sample Results (U/mL) |  |  |  |  |  |  |  |
| --- | --- | --- | --- | --- | --- | --- | --- |
| Level | % | Rep. 1 | Rep. 2 | Rep. 3 | Mean | Target | Bias |
| A | 0.0% | 0.0490 | 0.0833 | 0.0677 | 0.0667 | 0.0667 | NA |
| B | 10% | 15.3 | 16.3 | 15.5 | 15.7 | 13.4 | 4.4% |
| C | 20% | 27.6 | 32.7 | 30.2 | 30.2 | 26.6 | 3.9% |
| D | 30% | 42.2 | 43.8 | 46.8 | 44.3 | 39.9 | 2.9% |
| E | 40% | 56.7 | 56.8 | 56.9 | 56.8 | 53.2 | -0.3% |
| F | 50% | 73.6 | 66.0 | 71.8 | 70.5 | 66.5 | -0.7% |
| G | 60% | 82.8 | 83.6 | 89.9 | 85.4 | 79.8 | 0.6% |
| H | 70% | 98.0 | 99.2 | 91.3 | 96.2 | 93.0 | -2.8% |
| I | 80% | 115 | 111 | 118 | 115 | 106 | 1.5% |
| J | 90% | 119 | 120 | 120 | 120 | 120 | -5.7% |
| K | 100% | 143 | 157 | 142 | 147 | 133 | 4.6% |

Targets were set by linear regression using all levels.

**Table 8.** Additional DBS linearity results.

| Linearity Sample Results (U/mL) |  |  |  |  |  |  |  |
| --- | --- | --- | --- | --- | --- | --- | --- |
| Level | % | Rep. 1 | Rep. 2 | Rep. 3 | Mean | Target | Bias |
| A | 0% | 0.0840 | 0.0913 | 0.117 | 0.0974 | -3.07 | NA |
| B | 10% | 20.1 | 28.9 | 27.8 | 25.6 | 29.7 | -13.8% |
| C | 20% | 53.2 | 57.2 | 69.3 | 59.9 | 62.5 | -4.1% |
| D | 30% | 80.4 | 78.8 | 83.8 | 81 | 95.2 | -14.9% |
| E | 40% | 141 | 120 | 132 | 131 | 128 | 2.4% |
| F | 50% | 173 | 158 | 159 | 163 | 161 | 1.6% |
| G | 60% | 205 | 238 | 212 | 218 | 194 | 12.8% |
| H | 70% | 249 | 258* | 237 | 248 | 226 | 9.6% |
| I | 80% | 198 | 243 | 225 | 222 | 259 | -14.3% |
| J | 90% | 291* | 303* | 293* | 296 | 292 | 1.3% |
| K | 100% | 292* | 300* | 378* | 323 | 325 | -0.4% |

Targets were set by linear regression using all levels.
\*Extracts were manually diluted 10-fold and re-measured. Results were multiplied by 10.

### 5. DBS Inter-Assay Imprecision

Inter-assay imprecision was performed over 4 different days with two replicates being measured per run and two runs occurring per day for a total of 16 replicates from 6 different contrived blood samples. A single instrument was used for this study. DBS samples spanned an antibody concentration range of 0.517 (∼3 times DBS cutoff) to 177 U/mL (∼1000 times DBS cutoff) using two different reagent lots (53688601 and 54682501). Repeatability, within-laboratory imprecision, and total variance (% CV) was less than 20.0% (Table 9 and Table 10) for both reagent lots and is considered acceptable relative to FDA guidance for ligand binding assays (*3*). All the samples had total categorical agreement of 100.0%.

**Table 9.**
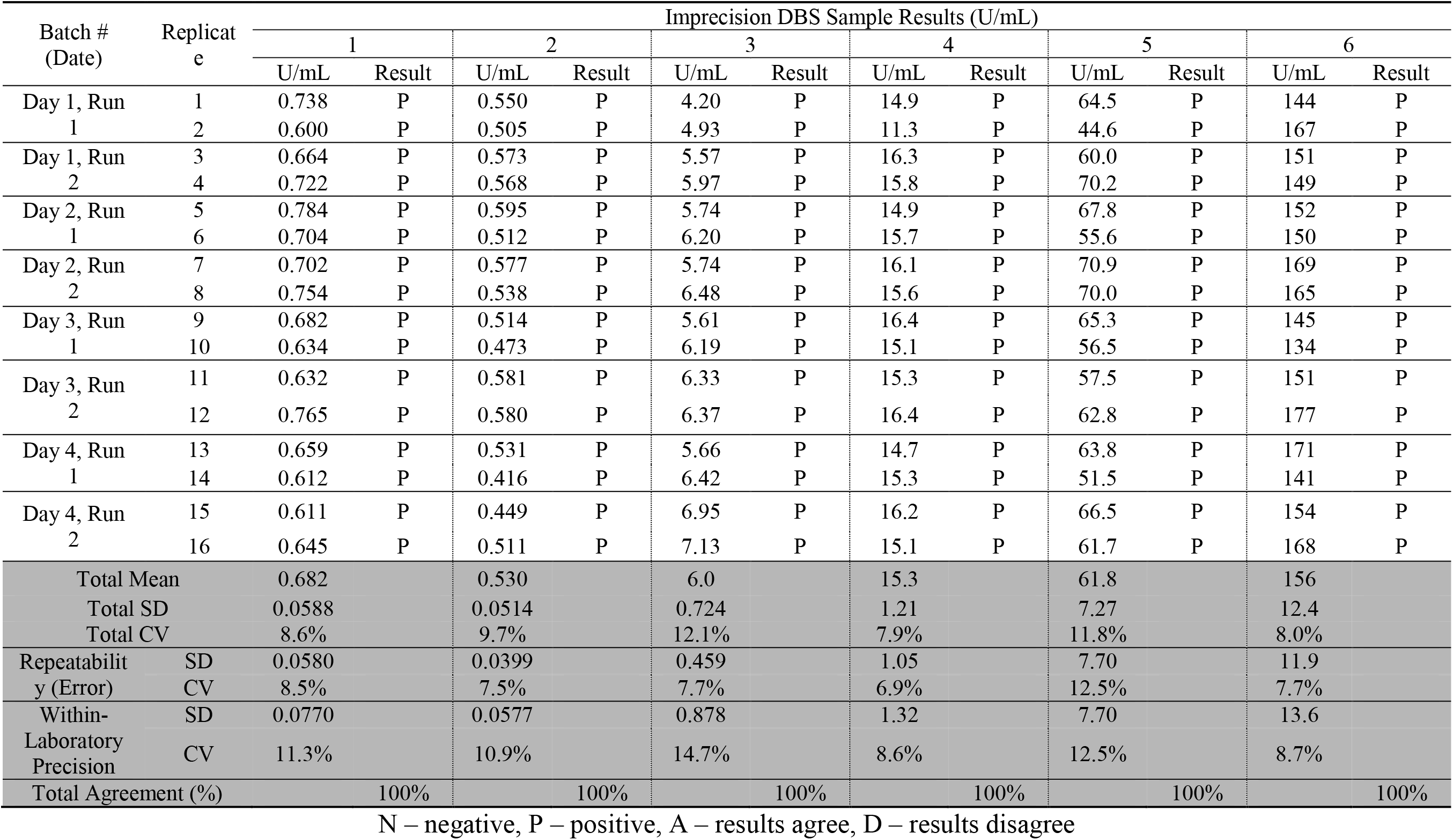
Inter-assay imprecision of extracted DBS samples (reagent lot 53688601)

**Table 10.**
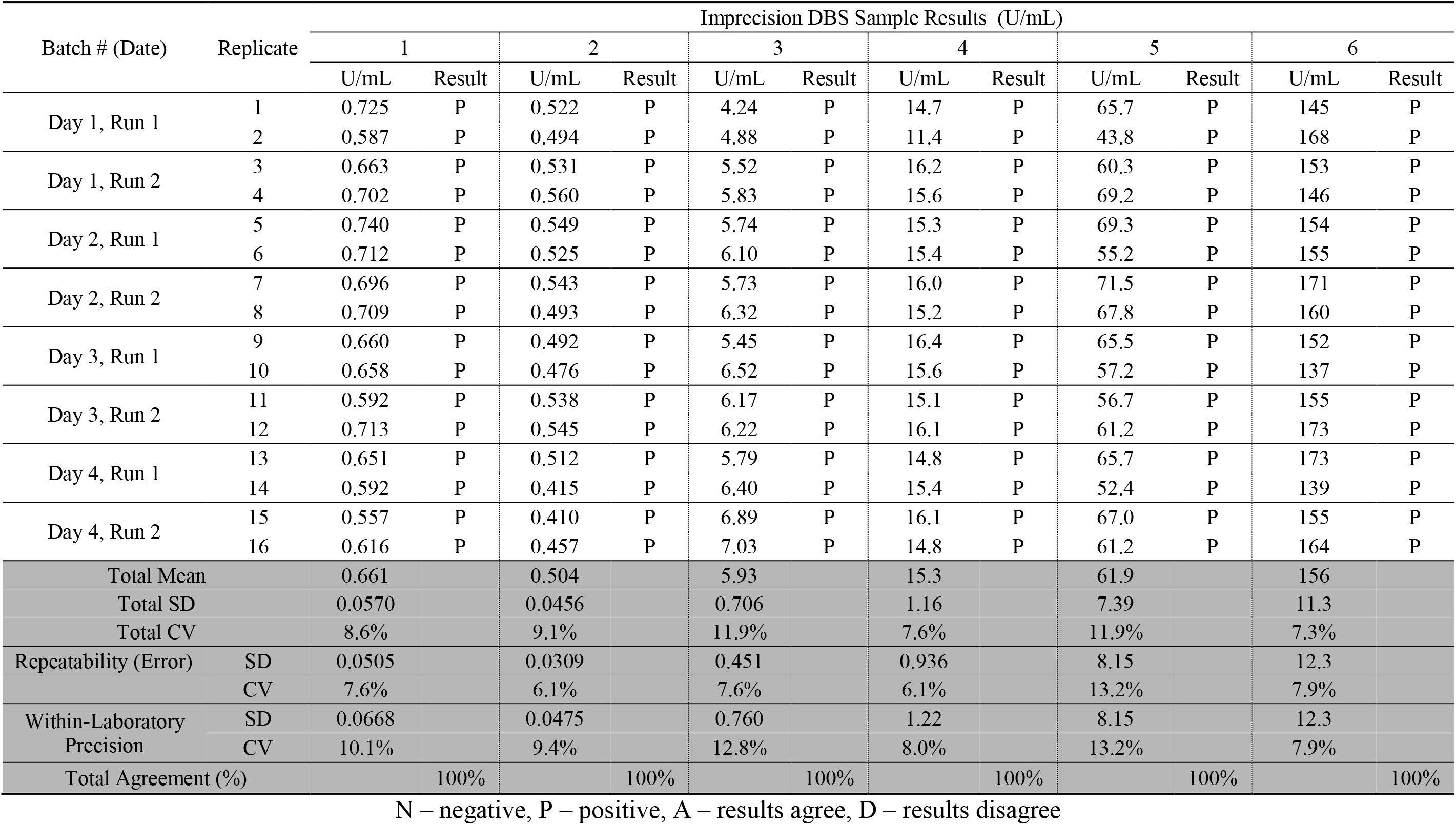
Inter-Assay imprecision of extracted DBS samples (reagent lot 54862501)

### 6. Clinical Cutoff

In order to report DBS samples categorically negative or positive, a clinical cutoff value was determined. This was calculated using the self-collected DBS samples from donors that were confirmed to be seronegative and did not have an active COVID-19 infection (Donors with “B” pre-fix in Table 11). The mean and standard deviation for results from self-collected DBS samples were found to be 0.0625 and 0.0405 U/mL, respectively (n = 78). The cutoff was set by multiplying the standard deviation by three and adding the mean to give a value of 0.184 U/mL. As such, the clinical cutoff was set to 0.185 U/mL where all results greater than or equal to this value were considered positive (P) and all values less than 0.185 considered negative (N).

### 7. Clinical Performance Study

For the clinical performance studies, some results were not obtained for all sample types from all donors (Table 11). Either a sample was not provided for testing or the sample did not have sufficient volume for measurement (*e.g.* insufficient volume on DBS card for extraction and measurement, QNS). Serum samples that initially measured greater than 250 U/mL were manually diluted 10-fold with the approved Roche Universal Diluent and re-measured; results were multiplied by 10. In addition, some samples had results that were reported by the instrument as “< Test” indicating the signal generated was less than the intercept of the calibration curve used to calculate a concentration. Donors with a pre-fix of “A” previously tested positive for COVID-19 using an RT-PCR test while donors with a pre-fix of “B” are presumed negative.

All serum and DBS results where a definitive number was not obtained were excluded from quantitative analysis. Six donors (B10, B35, B51, B64, B-077, B80) that were reported to have never had COVID-19 as well as confirmed not to have an active SARS-CoV-2 infection had venous serum, professionally collected DBS, and self-collected DBS samples results all measure as positive. To confirm these results, serum was measured using three additional EUA approved antibody assay: Roche Elecsys ® Anti-SARS-CoV-2 (qualitative nucleocapsid assay), DiaSorin Liason SARS-CoV-2 S1/S2 IgG (qualitative spike protein assay) and DiaSorin Laison SARS-CoV-2 IgM (qualitative spike protein assay) (*4*). All six donors had a positive serum test result for at least one additional assay (Table 12). As such, these donors were excluded from qualitative and quantitative method comparison data analysis.

**Table 11.**
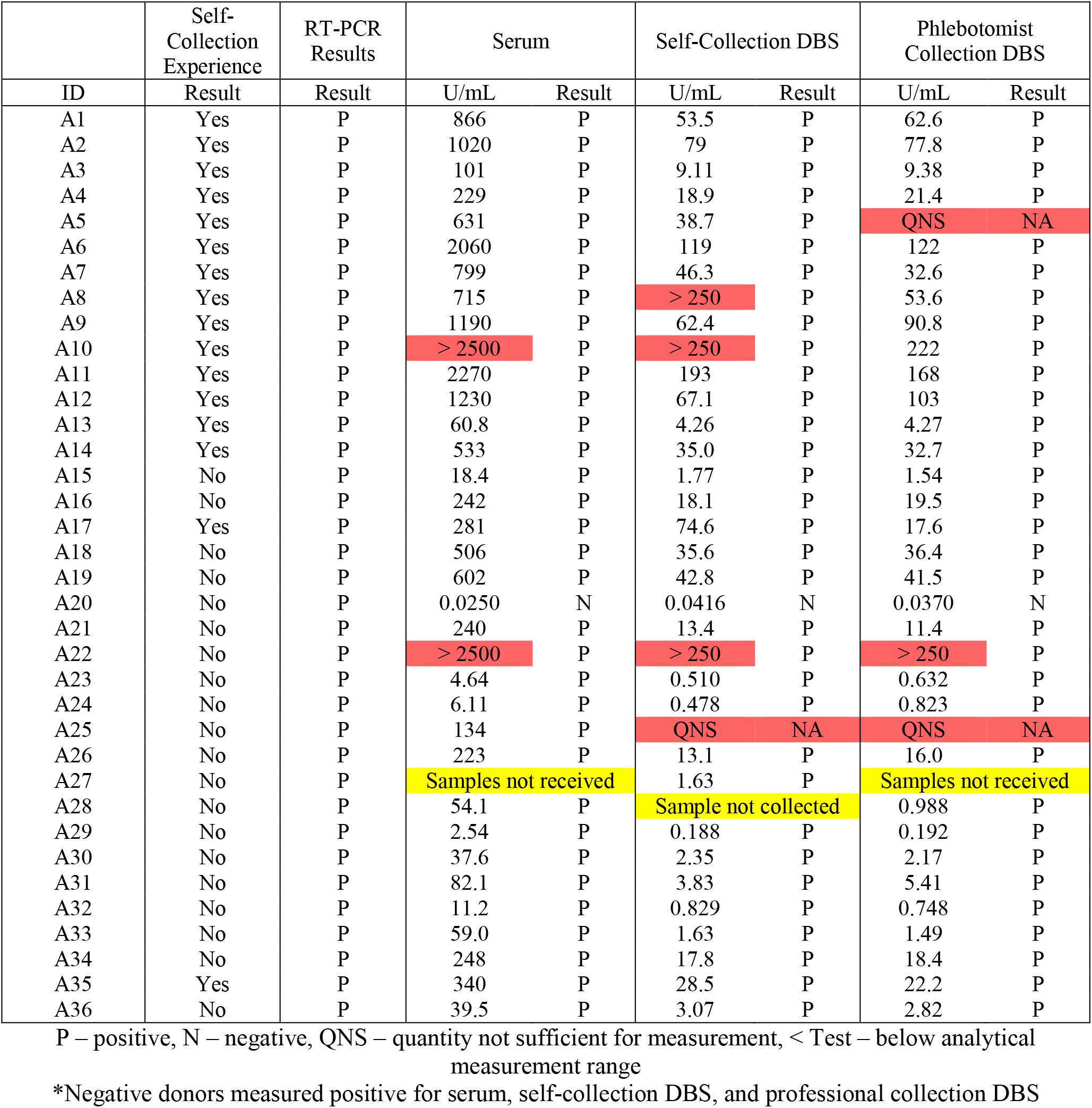
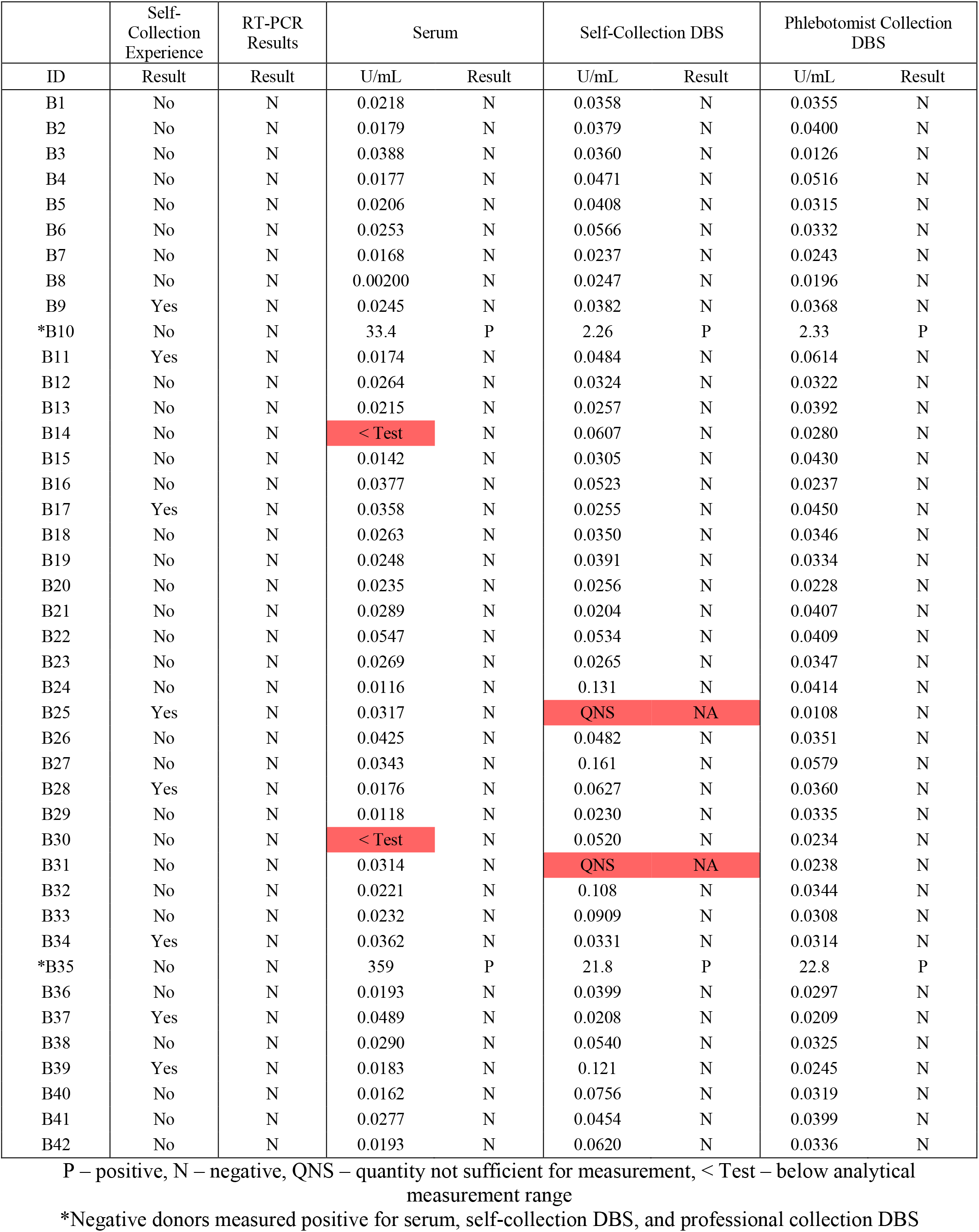
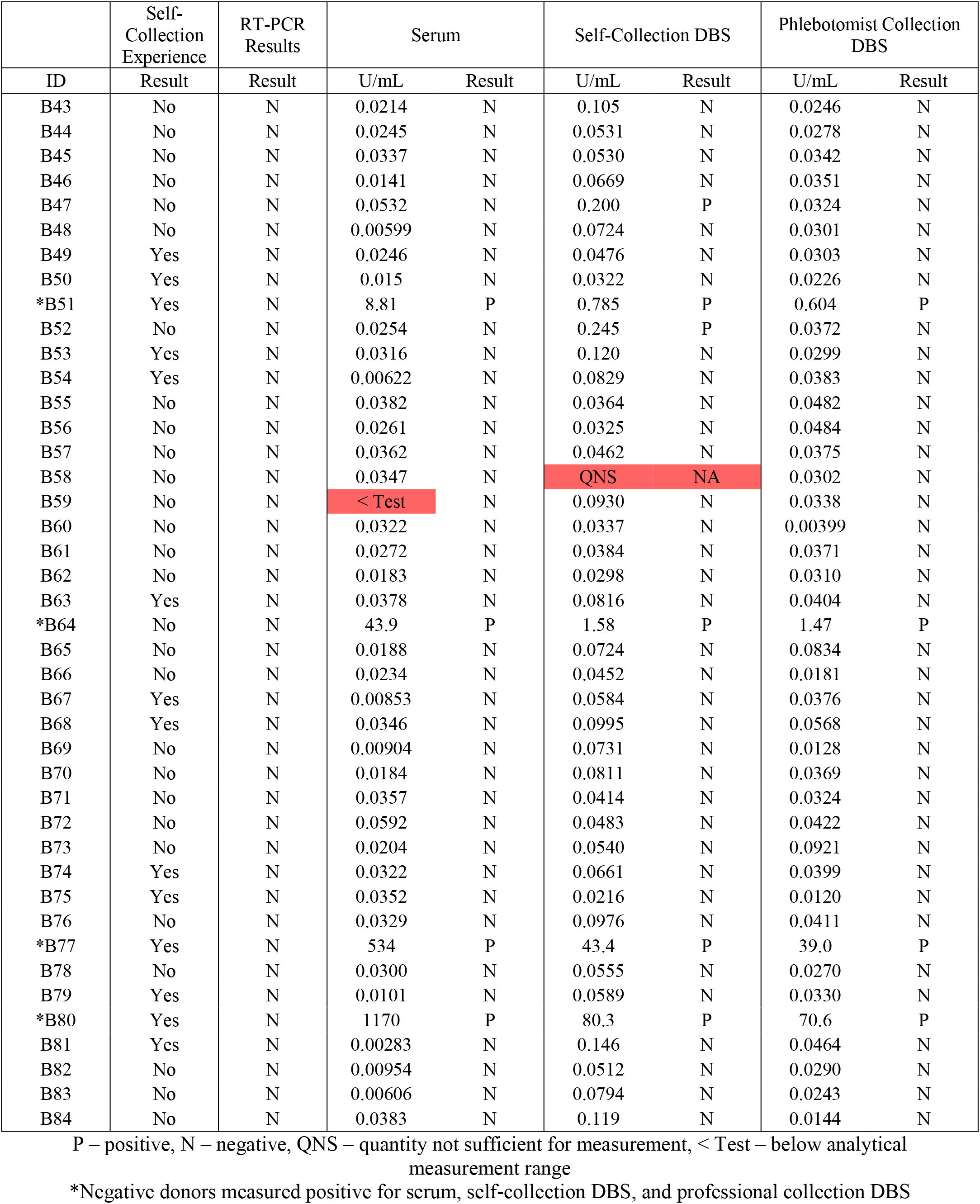
Clinical performance results.

|  | Self-Collection Experience | RT-PCR Results | Serum |  | Self-Collection DBS |  | Phlebotomist Collection DBS |  |
| --- | --- | --- | --- | --- | --- | --- | --- | --- |
| ID | Result | Result | U/mL | Result | U/mL | Result | U/mL | Result |
| A1 | Yes | P | 866 | P | 53.5 | P | 62.6 | P |
| A2 | Yes | P | 1020 | P | 79 | P | 77.8 | P |
| A3 | Yes | P | 101 | P | 9.11 | P | 9.38 | P |
| A4 | Yes | P | 229 | P | 18.9 | P | 21.4 | P |
| A5 | Yes | P | 631 | P | 38.7 | P | QNS | NA |
| A6 | Yes | P | 2060 | P | 119 | P | 122 | P |
| A7 | Yes | P | 799 | P | 46.3 | P | 32.6 | P |
| A8 | Yes | P | 715 | P | > 250 | P | 53.6 | P |
| A9 | Yes | P | 1190 | P | 62.4 | P | 90.8 | P |
| A10 | Yes | P | > 2500 | P | > 250 | P | 222 | P |
| A11 | Yes | P | 2270 | P | 193 | P | 168 | P |
| A12 | Yes | P | 1230 | P | 67.1 | P | 103 | P |
| A13 | Yes | P | 60.8 | P | 4.26 | P | 4.27 | P |
| A14 | Yes | P | 533 | P | 35.0 | P | 32.7 | P |
| A15 | No | P | 18.4 | P | 1.77 | P | 1.54 | P |
| A16 | No | P | 242 | P | 18.1 | P | 19.5 | P |
| A17 | Yes | P | 281 | P | 74.6 | P | 17.6 | P |
| A18 | No | P | 506 | P | 35.6 | P | 36.4 | P |
| A19 | No | P | 602 | P | 42.8 | P | 41.5 | P |
| A20 | No | P | 0.0250 | N | 0.0416 | N | 0.0370 | N |
| A21 | No | P | 240 | P | 13.4 | P | 11.4 | P |
| A22 | No | P | > 2500 | P | > 250 | P | > 250 | P |
| A23 | No | P | 4.64 | P | 0.510 | P | 0.632 | P |
| A24 | No | P | 6.11 | P | 0.478 | P | 0.823 | P |
| A25 | No | P | 134 | P | QNS | NA | QNS | NA |
| A26 | No | P | 223 | P | 13.1 | P | 16.0 | P |
| A27 | No | P | Samples not received |  | 1.63 | P | Samples not received |  |
| A28 | No | P | 54.1 | P | Sample not collected |  | 0.988 | P |
| A29 | No | P | 2.54 | P | 0.188 | P | 0.192 | P |
| A30 | No | P | 37.6 | P | 2.35 | P | 2.17 | P |
| A31 | No | P | 82.1 | P | 3.83 | P | 5.41 | P |
| A32 | No | P | 11.2 | P | 0.829 | P | 0.748 | P |
| A33 | No | P | 59.0 | P | 1.63 | P | 1.49 | P |
| A34 | No | P | 248 | P | 17.8 | P | 18.4 | P |
| A35 | Yes | P | 340 | P | 28.5 | P | 22.2 | P |
| A36 | No | P | 39.5 | P | 3.07 | P | 2.82 | P |
| ID | Result | Result | U/mL | Result | U/mL | Result | U/mL | Result |
| B1 | No | N | 0.0218 | N | 0.0358 | N | 0.0355 | N |
| B2 | No | N | 0.0179 | N | 0.0379 | N | 0.0400 | N |
| B3 | No | N | 0.0388 | N | 0.0360 | N | 0.0126 | N |
| B4 | No | N | 0.0177 | N | 0.0471 | N | 0.0516 | N |
| B5 | No | N | 0.0206 | N | 0.0408 | N | 0.0315 | N |
| B6 | No | N | 0.0253 | N | 0.0566 | N | 0.0332 | N |
| B7 | No | N | 0.0168 | N | 0.0237 | N | 0.0243 | N |
| B8 | No | N | 0.00200 | N | 0.0247 | N | 0.0196 | N |
| B9 | Yes | N | 0.0245 | N | 0.0382 | N | 0.0368 | N |
| *B10 | No | N | 33.4 | P | 2.26 | P | 2.33 | P |
| B11 | Yes | N | 0.0174 | N | 0.0484 | N | 0.0614 | N |
| B12 | No | N | 0.0264 | N | 0.0324 | N | 0.0322 | N |
| B13 | No | N | 0.0215 | N | 0.0257 | N | 0.0392 | N |
| B14 | No | N | < Test | N | 0.0607 | N | 0.0280 | N |
| B15 | No | N | 0.0142 | N | 0.0305 | N | 0.0430 | N |
| B16 | No | N | 0.0377 | N | 0.0523 | N | 0.0237 | N |
| B17 | Yes | N | 0.0358 | N | 0.0255 | N | 0.0450 | N |
| B18 | No | N | 0.0263 | N | 0.0350 | N | 0.0346 | N |
| B19 | No | N | 0.0248 | N | 0.0391 | N | 0.0334 | N |
| B20 | No | N | 0.0235 | N | 0.0256 | N | 0.0228 | N |
| B21 | No | N | 0.0289 | N | 0.0204 | N | 0.0407 | N |
| B22 | No | N | 0.0547 | N | 0.0534 | N | 0.0409 | N |
| B23 | No | N | 0.0269 | N | 0.0265 | N | 0.0347 | N |
| B24 | No | N | 0.0116 | N | 0.131 | N | 0.0414 | N |
| B25 | Yes | N | 0.0317 | N | QNS | NA | 0.0108 | N |
| B26 | No | N | 0.0425 | N | 0.0482 | N | 0.0351 | N |
| B27 | No | N | 0.0343 | N | 0.161 | N | 0.0579 | N |
| B28 | Yes | N | 0.0176 | N | 0.0627 | N | 0.0360 | N |
| B29 | No | N | 0.0118 | N | 0.0230 | N | 0.0335 | N |
| B30 | No | N | < Test | N | 0.0520 | N | 0.0234 | N |
| B31 | No | N | 0.0314 | N | QNS | NA | 0.0238 | N |
| B32 | No | N | 0.0221 | N | 0.108 | N | 0.0344 | N |
| B33 | No | N | 0.0232 | N | 0.0909 | N | 0.0308 | N |
| B34 | Yes | N | 0.0362 | N | 0.0331 | N | 0.0314 | N |
| *B35 | No | N | 359 | P | 21.8 | P | 22.8 | P |
| B36 | No | N | 0.0193 | N | 0.0399 | N | 0.0297 | N |
| B37 | Yes | N | 0.0489 | N | 0.0208 | N | 0.0209 | N |
| B38 | No | N | 0.0290 | N | 0.0540 | N | 0.0325 | N |
| B39 | Yes | N | 0.0183 | N | 0.121 | N | 0.0245 | N |
| B40 | No | N | 0.0162 | N | 0.0756 | N | 0.0319 | N |
| B41 | No | N | 0.0277 | N | 0.0454 | N | 0.0399 | N |
| B42 | No | N | 0.0193 | N | 0.0620 | N | 0.0336 | N |
| ID | Result | Result | U/mL | Result | U/mL | Result | U/mL | Result |
| B43 | No | N | 0.0214 | N | 0.105 | N | 0.0246 | N |
| B44 | No | N | 0.0245 | N | 0.0531 | N | 0.0278 | N |
| B45 | No | N | 0.0337 | N | 0.0530 | N | 0.0342 | N |
| B46 | No | N | 0.0141 | N | 0.0669 | N | 0.0351 | N |
| B47 | No | N | 0.0532 | N | 0.200 | P | 0.0324 | N |
| B48 | No | N | 0.00599 | N | 0.0724 | N | 0.0301 | N |
| B49 | Yes | N | 0.0246 | N | 0.0476 | N | 0.0303 | N |
| B50 | Yes | N | 0.015 | N | 0.0322 | N | 0.0226 | N |
| *B51 | Yes | N | 8.81 | P | 0.785 | P | 0.604 | P |
| B52 | No | N | 0.0254 | N | 0.245 | P | 0.0372 | N |
| B53 | Yes | N | 0.0316 | N | 0.120 | N | 0.0299 | N |
| B54 | Yes | N | 0.00622 | N | 0.0829 | N | 0.0383 | N |
| B55 | No | N | 0.0382 | N | 0.0364 | N | 0.0482 | N |
| B56 | No | N | 0.0261 | N | 0.0325 | N | 0.0484 | N |
| B57 | No | N | 0.0362 | N | 0.0462 | N | 0.0375 | N |
| B58 | No | N | 0.0347 | N | QNS | NA | 0.0302 | N |
| B59 | No | N | < Test | N | 0.0930 | N | 0.0338 | N |
| B60 | No | N | 0.0322 | N | 0.0337 | N | 0.00399 | N |
| B61 | No | N | 0.0272 | N | 0.0384 | N | 0.0371 | N |
| B62 | No | N | 0.0183 | N | 0.0298 | N | 0.0310 | N |
| B63 | Yes | N | 0.0378 | N | 0.0816 | N | 0.0404 | N |
| *B64 | No | N | 43.9 | P | 1.58 | P | 1.47 | P |
| B65 | No | N | 0.0188 | N | 0.0724 | N | 0.0834 | N |
| B66 | No | N | 0.0234 | N | 0.0452 | N | 0.0181 | N |
| B67 | Yes | N | 0.00853 | N | 0.0584 | N | 0.0376 | N |
| B68 | Yes | N | 0.0346 | N | 0.0995 | N | 0.0568 | N |
| B69 | No | N | 0.00904 | N | 0.0731 | N | 0.0128 | N |
| B70 | No | N | 0.0184 | N | 0.0811 | N | 0.0369 | N |
| B71 | No | N | 0.0357 | N | 0.0414 | N | 0.0324 | N |
| B72 | No | N | 0.0592 | N | 0.0483 | N | 0.0422 | N |
| B73 | No | N | 0.0204 | N | 0.0540 | N | 0.0921 | N |
| B74 | Yes | N | 0.0322 | N | 0.0661 | N | 0.0399 | N |
| B75 | Yes | N | 0.0352 | N | 0.0216 | N | 0.0120 | N |
| B76 | No | N | 0.0329 | N | 0.0976 | N | 0.0411 | N |
| *B77 | Yes | N | 534 | P | 43.4 | P | 39.0 | P |
| B78 | No | N | 0.0300 | N | 0.0555 | N | 0.0270 | N |
| B79 | Yes | N | 0.0101 | N | 0.0589 | N | 0.0330 | N |
| *B80 | Yes | N | 1170 | P | 80.3 | P | 70.6 | P |
| B81 | Yes | N | 0.00283 | N | 0.146 | N | 0.0464 | N |
| B82 | No | N | 0.00954 | N | 0.0512 | N | 0.0290 | N |
| B83 | No | N | 0.00606 | N | 0.0794 | N | 0.0243 | N |
| B84 | No | N | 0.0383 | N | 0.119 | N | 0.0144 | N |

P – positive, N – negative, QNS – quantity not sufficient for measurement, < Test – below analytical measurement range
\*Negative donors measured positive for serum, self-collection DBS, and professional collection DBS

**Table 12:** Six “false” positive donor serum results from four EUA approved serological assays.

|  |  | EUA Approved Assay |  |  |  |  |  |  |  |
| --- | --- | --- | --- | --- | --- | --- | --- | --- | --- |
|  |  | Roche Elecsys<br>Anti-SARS-CoV-2 S |  | Roche Elecsys<br>Anti-SARS-CoV-2 |  | DiaSorin Liaison<br>SARS-CoV-2 S1/S2<br>IgG |  | DiaSorin Liaison<br>SARS-CoV-2<br>IgM |  |
| Target<br>Assay Type |  | Spike Protein<br>Semi-Quantitative |  | Nucleocapsid<br>Qualitative |  | Spike Protein<br>Qualitative |  | Spike Protein<br>Qualitative |  |
| Negative Range |  | < 0.8 U/mL |  | < 1.0 COI |  | < 12 AU/mL |  | < 1.1 index |  |
| Equivocal Range |  | not applicable |  | not applicable |  | 12 to 15 AU/mL |  | not applicable |  |
| Positive Range |  | ≥ 0.8 U/mL |  | ≥ 1.0 COI |  | ≥ 15 AU/mL |  | ≥ 1.1 index |  |
|  |  | Result<br>(U/mL) | Interpret. | Result<br>(COI) | Interpret. | Result<br>(AU/mL) | Interpret. | Result<br>(index) | Interpret. |
| Donor ID | B10 | 33.4 | P | 10.85 | P | 10.5 | N | 0.452 | N |
|  | B35 | 359 | P | 55.63 | P | 48 | P | 0.15 | N |
|  | B51 | 8.81 | P | 0.625 | N | 8.22 | N | 1.49 | P |
|  | B64 | 43.9 | P | 125.7 | P | 50.6 | P | 0.905 | N |
|  | B77 | 534 | P | 10.2 | P | 111 | P | 0.39 | N |
|  | B80 | 1170 | P | 0.089 | N | 233 | P | 0.132 | N |

### 8. DBS Shipping Stability

Studies were performed to evaluate sample stability during the shipping process including storage of samples before shipping, samples sitting prior to pick up, and conditions following pick up and transport to the lab for testing (*5,6*). Contrived blood samples (n = 76) were prepared with antibody concentrations ranging from 0.0358 U/mL (∼0.2 times DBS cutoff) to 4.91 U/mL (∼27 times DBS cutoff); most samples (n = 70) were within 5 times the DBS cutoff. Samples were made into triplicates and were divided into three shipping conditions: ambient (20-25°C), winter, and summer (refer to Table 13). Samples were measured in parallel whereas the room temperature samples were used as baseline results. Results that were less than the assay’s DBS LOQ were excluded from bias analysis but included in qualitative analysis. Winter and summer excursion samples had a total categorical agreement of 97.4% and mean biases of 6.0% and −0.5%, respectively. One winter excursion sample that did not have categorical agreement had a baseline concentration of 0.0499 U/mL and a concentration post-excursion of 0.269 U/mL which is within 1.5 times the DBS clinical cutoff (0.185 U/mL). The other winter-excursion sample that did not have categorical agreement with baseline results had a baseline concentration of 0.185 U/mL which is equivalent to the DBS clinical cutoff (0.185 U/mL). The two summer-excursion samples that did not have categorical agreement with baseline results included this sample with baseline results equivalent to the DBS clinical cutoff and a sample with baseline results of 0.233 U/mL which is within 26% of the DBS clinical cutoff.

**Table 13.**
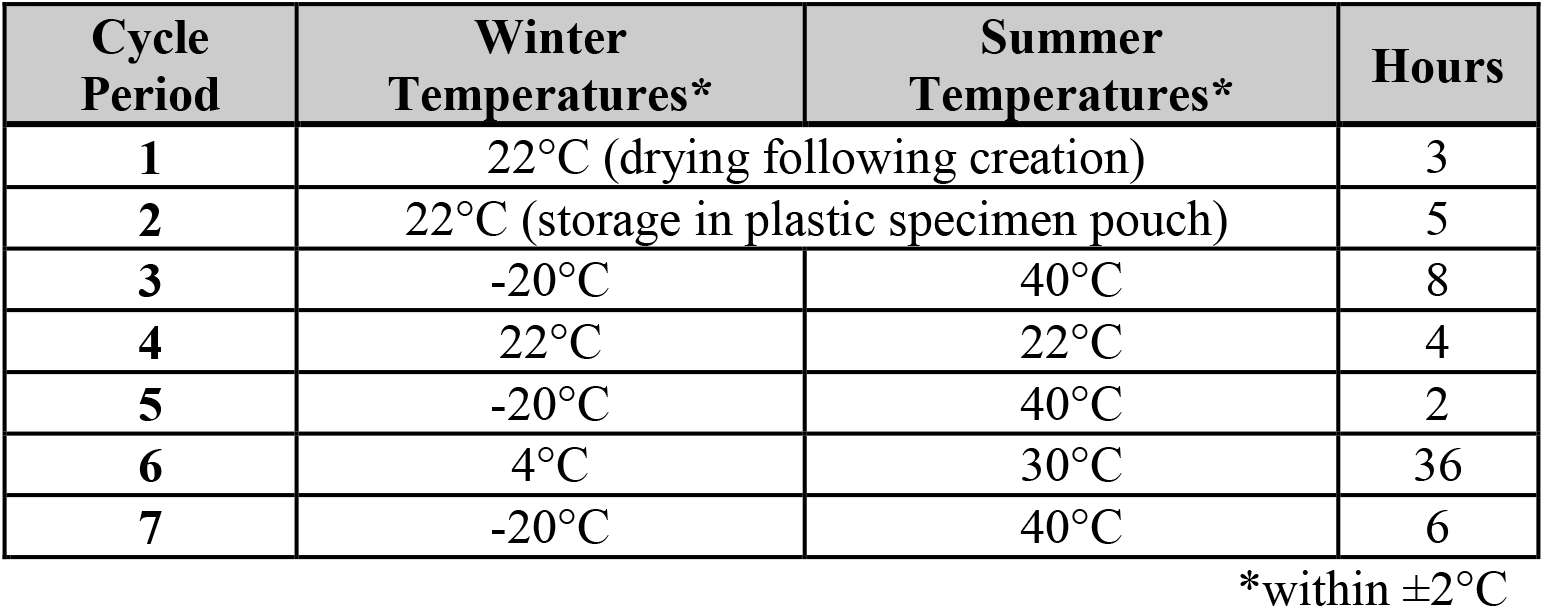
Simulated shipping conditions.

**Table 14.** DBS shipping excursion results.

| Sample | Ambient |  | Winter Excursion |  |  |  | Summer Excursion |  |  |  |
| --- | --- | --- | --- | --- | --- | --- | --- | --- | --- | --- |
|  | U/mL | Result | U/mL | Bias | Result | Agree / Disagree | U/mL | Bias | Result | Agree / Disagree |
| 1 | 0.0358 | N | 0.0624 | NA | N | A | 0.140 | NA | N | A |
| 2 | 0.0446 | N | 0.119 | NA | N | A | 0.0612 | NA | N | A |
| 3 | 0.0455 | N | 0.0469 | NA | N | A | 0.0650 | NA | N | A |
| 4 | 0.0499 | N | 0.269 | NA | P | D | 0.0978 | NA | N | A |
| 5 | 0.0534 | N | 0.102 | NA | N | A | 0.0681 | NA | N | A |
| 6 | 0.0641 | N | 0.0370 | NA | N | A | 0.0288 | NA | N | A |
| 7 | 0.0663 | N | 0.0384 | NA | N | A | 0.0473 | NA | N | A |
| 8 | 0.0800 | N | 0.0390 | NA | N | A | 0.0434 | NA | N | A |
| 9 | 0.0966 | N | 0.0398 | NA | N | A | 0.0232 | NA | N | A |
| 10 | 0.111 | N | 0.0405 | NA | N | A | 0.0281 | NA | N | A |
| 11 | 0.113 | N | 0.109 | NA | N | A | 0.112 | NA | N | A |
| 12 | 0.123 | N | 0.137 | NA | N | A | 0.148 | NA | N | A |
| 13 | 0.151 | N | 0.171 | NA | N | A | 0.142 | NA | N | A |
| 14 | 0.164 | N | 0.183 | NA | N | A | 0.121 | NA | N | A |
| 15 | 0.169 | N | 0.173 | NA | N | A | 0.177 | NA | N | A |
| 16 | 0.176 | N | 0.155 | NA | N | A | 0.138 | NA | N | A |
| 17 | 0.177 | N | 0.181 | NA | N | A | 0.147 | NA | N | A |
| 18 | 0.185 | P | 0.174 | NA | N | D | 0.144 | NA | N | D |
| 19 | 0.195 | P | 0.497 | 154.9% | P | A | 0.242 | 24.1% | P | A |
| 20 | 0.205 | P | 0.237 | 15.6% | P | A | 0.220 | 7.3% | P | A |
| 21 | 0.205 | P | 0.232 | 13.2% | P | A | 0.220 | 7.3% | P | A |
| 22 | 0.212 | P | 0.218 | 2.8% | P | A | 0.236 | 11.3% | P | A |
| 23 | 0.233 | P | 0.209 | -10.3% | P | A | 0.165 | NA | N | D |
| 24 | 0.250 | P | 0.206 | -17.6% | P | A | 0.194 | -22.4% | P | A |
| 25 | 0.259 | P | 0.235 | -9.3% | P | A | 0.235 | -9.3% | P | A |
| 26 | 0.259 | P | 0.221 | -14.7% | P | A | 0.196 | -24.3% | P | A |
| 27 | 0.262 | P | 0.223 | -14.9% | P | A | 0.242 | -7.6% | P | A |
| 28 | 0.277 | P | 0.259 | -6.5% | P | A | 0.230 | -17.0% | P | A |
| 29 | 0.304 | P | 0.242 | -20.4% | P | A | 0.284 | -6.6% | P | A |
| 30 | 0.322 | P | 0.248 | -23.0% | P | A | 0.318 | -1.2% | P | A |
| 31 | 0.330 | P | 0.385 | 16.7% | P | A | 0.318 | -3.6% | P | A |
| 32 | 0.332 | P | 0.362 | 9.0% | P | A | 0.328 | -1.2% | P | A |
| 33 | 0.334 | P | 0.322 | -3.6% | P | A | 0.360 | 7.8% | P | A |
| 34 | 0.344 | P | 0.422 | 22.7% | P | A | 0.362 | 5.2% | P | A |
| 35 | 0.357 | P | 0.409 | 14.6% | P | A | 0.352 | -1.4% | P | A |
| 36 | 0.359 | P | 0.349 | -2.8% | P | A | 0.386 | 7.5% | P | A |
| 37 | 0.360 | P | 0.458 | 27.2% | P | A | 0.369 | 2.5% | P | A |
| 38 | 0.360 | P | 0.430 | 19.4% | P | A | 0.397 | 10.3% | P | A |

N – negative, P – positive, A – results agree, D – results disagree
| Sample | Ambient |  | Winter Excursion |  |  |  | Summer Excursion |  |  |  |
| --- | --- | --- | --- | --- | --- | --- | --- | --- | --- | --- |
|  | U/mL | Result | U/mL | Bias | Result | Agree /<br>Disagree | U/mL | Bias | Result | Agree /<br>Disagree |
| 39 | 0.365 | P | 0.353 | -3.3% | P | A | 0.357 | -2.2% | P | A |
| 40 | 0.365 | P | 0.473 | 29.6% | P | A | 0.401 | 9.9% | P | A |
| 41 | 0.366 | P | 0.399 | 9.0% | P | A | 0.345 | -5.7% | P | A |
| 42 | 0.371 | P | 0.393 | 5.9% | P | A | 0.376 | 1.3% | P | A |
| 43 | 0.372 | P | 0.435 | 16.9% | P | A | 0.397 | 6.7% | P | A |
| 44 | 0.386 | P | 0.439 | 13.7% | P | A | 0.424 | 9.8% | P | A |
| 45 | 0.394 | P | 0.427 | 8.4% | P | A | 0.403 | 2.3% | P | A |
| 46 | 0.394 | P | 0.425 | 7.9% | P | A | 0.404 | 2.5% | P | A |
| 47 | 0.394 | P | 0.437 | 10.9% | P | A | 0.387 | -1.8% | P | A |
| 48 | 0.402 | P | 0.416 | 3.5% | P | A | 0.405 | 0.7% | P | A |
| 49 | 0.405 | P | 0.443 | 9.4% | P | A | 0.455 | 12.3% | P | A |
| 50 | 0.414 | P | 0.496 | 19.8% | P | A | 0.445 | 7.5% | P | A |
| 51 | 0.422 | P | 0.436 | 3.3% | P | A | 0.431 | 2.1% | P | A |
| 52 | 0.439 | P | 0.439 | 0.0% | P | A | 0.436 | -0.7% | P | A |
| 53 | 0.456 | P | 0.464 | 1.8% | P | A | 0.445 | -2.4% | P | A |
| 54 | 0.474 | P | 0.492 | 3.8% | P | A | 0.441 | -7.0% | P | A |
| 55 | 0.486 | P | 0.496 | 2.1% | P | A | 0.491 | 1.0% | P | A |
| 56 | 0.488 | P | 0.619 | 26.8% | P | A | 0.529 | 8.4% | P | A |
| 57 | 0.499 | P | 0.502 | 0.6% | P | A | 0.422 | -15.4% | P | A |
| 58 | 0.551 | P | 0.532 | -3.4% | P | A | 0.588 | 6.7% | P | A |
| 59 | 0.672 | P | 0.657 | -2.2% | P | A | 0.743 | 10.6% | P | A |
| 60 | 0.705 | P | 0.736 | 4.4% | P | A | 0.718 | 1.8% | P | A |
| 61 | 0.714 | P | 0.730 | 2.8% | P | A | 0.654 | -8.4% | P | A |
| 62 | 0.752 | P | 0.759 | 0.9% | P | A | 0.641 | -14.8% | P | A |
| 63 | 0.811 | P | 0.750 | -7.5% | P | A | 0.803 | -1.0% | P | A |
| 64 | 0.812 | P | 0.929 | 14.4% | P | A | 0.822 | 1.2% | P | A |
| 65 | 0.900 | P | 0.980 | 8.9% | P | A | 0.886 | -1.6% | P | A |
| 66 | 0.902 | P | 0.937 | 3.9% | P | A | 0.893 | -1.0% | P | A |
| 67 | 0.981 | P | 0.988 | 0.7% | P | A | 1.04 | 6.0% | P | A |
| 68 | 1.04 | P | 0.930 | -10.8% | P | A | 0.954 | -8.3% | P | A |
| 69 | 1.05 | P | 1.00 | -4.8% | P | A | 0.970 | -7.6% | P | A |
| 70 | 1.17 | P | 1.10 | -6.0% | P | A | 1.23 | 5.1% | P | A |
| 71 | 1.28 | P | 1.35 | 5.5% | P | A | 1.25 | -2.3% | P | A |
| 72 | 1.52 | P | 1.60 | 5.3% | P | A | 1.57 | 3.3% | P | A |
| 73 | 1.53 | P | 1.62 | 5.9% | P | A | 1.40 | -8.5% | P | A |
| 74 | 1.71 | P | 1.61 | -5.8% | P | A | 1.15 | -32.7% | P | A |
| 75 | 2.31 | P | 2.27 | -1.7% | P | A | 2.24 | -3.0% | P | A |
| 76 | 4.68 | P | 4.30 | -8.1% | P | A | 4.91 | 4.9% | P | A |
|  |  |  | Mean Bias: | 6.0% | Total Agreement: | 97.4% | Mean Bias | -0.5% | Total Agreement: | 97.4% |
|  |  |  | n | 58 | n | 76 | n | 57 | n | 76 |

### 9. Robustness Studies

Additional robustness studies were performed to stress different aspects of sample collection. Contrived blood samples were prepared with antibody concentrations ranging from 0.0380 U/mL (∼0.2 times DBS cutoff) to 1.52 U/mL (∼8 times DBS cutoff); most samples were within 5 times the DBS cutoff. All samples used to generate baseline results were spotted and dried for three hours prior to storage; no interferents or contamination was introduced. For acceptance of results, biases were determined to baseline measurements for results greater than the assay’s LOQ. A mean bias of 20% was used as quantitative acceptance following FDA guidance for ligand binding assays (*3*). For qualitative assessment, a total categorical agreement of 95.0% was utilized based on guidance from the FDA’s Home Specimen Collection Serology Template (*5*).

#### 9.1. Alternate Drying Times

To test the effects of drying time on sample results, alternative drying times were investigated using antibody concentrations designed to stress the clinical cutoff. Drying times prior to storage were 0, 1, 3 (baseline), and 22 hours (overnight). Following the pre-determined drying time, samples were placed in plastic specimen pouches (without desiccant) and stored at room temperature until measurement. DBS samples that were immediately stored after being spotted demonstrated total categorical agreement of 95.0% and mean bias of −32.5% and total categorical agreement of 95.0% versus DBS samples stored for 3 hours prior to placing in a desiccant free specimen pouch. The one sample that did not have categorical agreement with baseline results had a baseline concentration of 0.193 U/mL which is within 5% of the DBS clinical cutoff (0.185 U/mL). DBS samples that dried for one hour demonstrated total categorical agreement of 95.0% and mean bias of 1.3%. The one sample that did not have categorical agreement with baseline results had a baseline concentration of 0.176 U/mL which is again within 5% of the DBS clinical cutoff (0.185 U/mL). DBS samples stored for 22 hours demonstrated total categorical agreement of 100% and mean bias of 1.3% (Table 15). These results indicate that at least 1 hour of drying is required prior to sample storage and shipment. Risk of donors not allowing sufficient drying can be mitigated by appropriate design of donor materials including instructions for use (IFUs) and demonstration videos.

**Table 15.** Alternative drying times of 0, 1, 3 (baseline) and 22 hours for DBS samples.

| Sample | Baseline (3 Hrs) |  | Drying Time Prior to Storage (Hrs) |  |  |  |  |  |  |  |  |  |  |  |
| --- | --- | --- | --- | --- | --- | --- | --- | --- | --- | --- | --- | --- | --- | --- |
|  |  |  | 0 Hrs |  |  |  | 1 Hr |  |  |  | 22 Hrs |  |  |  |
|  | U/mL | Result | U/mL | Bias | Result | Agree/<br>Disagree | U/mL | Bias | Result | Agree/<br>Disagree | U/mL | Bias | Result | Agree/<br>Disagree |
| 1 | 0.0380 | N | 0.0594 | NA | N | A | 0.0686 | NA | N | A | 0.0553 | NA | N | A |
| 2 | 0.0439 | N | 0.0396 | NA | N | A | 0.0456 | NA | N | A | 0.0346 | NA | N | A |
| 3 | 0.0465 | N | 0.0770 | NA | N | A | 0.0668 | NA | N | A | 0.0255 | NA | N | A |
| 4 | 0.0512 | N | 0.0427 | NA | N | A | 0.0258 | NA | N | A | 0.0430 | NA | N | A |
| 5 | 0.0803 | N | 0.0491 | NA | N | A | 0.130 | NA | N | A | 0.0568 | NA | N | A |
| 6 | 0.166 | N | 0.144 | NA | N | A | 0.177 | NA | N | A | 0.170 | NA | N | A |
| 7 | 0.176 | N | 0.147 | NA | N | A | 0.233 | NA | P | D | 0.164 | NA | N | A |
| 8 | 0.193 | P | 0.136 | NA | N | D | 0.204 | 5.7% | P | A | 0.201 | 5.7% | P | A |
| 9 | 0.361 | P | 0.281 | -22.2% | P | A | 0.376 | 4.2% | P | A | 0.366 | 4.2% | P | A |
| 10 | 0.410 | P | 0.295 | -28.0% | P | A | 0.411 | 0.20% | P | A | 0.435 | 0.2% | P | A |
| 11 | 0.488 | P | 0.336 | -31.1% | P | A | 0.511 | 4.7% | P | A | 0.495 | 4.7% | P | A |
| 12 | 0.657 | P | 0.395 | -39.9% | P | A | 0.623 | -5.2% | P | A | 0.595 | -5.2% | P | A |
| 13 | 0.684 | P | 0.467 | -31.7% | P | A | 0.630 | -7.9% | P | A | 0.645 | -7.9% | P | A |
| 14 | 0.837 | P | 0.623 | -25.6% | P | A | 0.890 | 6.3% | P | A | 0.933 | 6.3% | P | A |
| 15 | 0.926 | P | 0.674 | -27.2% | P | A | 0.987 | 6.6% | P | A | 0.821 | 6.6% | P | A |
| 16 | 0.936 | P | 0.579 | -38.1% | P | A | 0.907 | -3.1% | P | A | 0.946 | -3.1% | P | A |
| 17 | 0.987 | P | 0.594 | -39.8% | P | A | 0.830 | -15.9% | P | A | 0.811 | -15.9% | P | A |
| 18 | 1.36 | P | 0.839 | -38.3% | P | A | 1.29 | -5.1% | P | A | 1.46 | -5.1% | P | A |
| 19 | 1.48 | P | 1.06 | -28.4% | P | A | 1.49 | 0.7% | P | A | 1.45 | 0.7% | P | A |
| 20 | 1.52 | P | 0.919 | -39.5% | P | A | 1.40 | -7.9% | P | A | 1.39 | -7.9% | P | A |
|  |  |  | Mean Bias | -32.5% | Total Agreement: | 95.0% | Mean Bias | 1.3% | Total Agreement: | 95.0% | Mean Bias | 1.3% | Total Agreement: | 100.0% |

N – negative, P – positive, A – results agree, D – results disagree

#### 9.2. DBS Card Drying in Humid Environment

Studies were performed to evaluate the effects of DBS samples drying under humid conditions. Four sample replicates were created for each concentration, where 3 of the 4 DBS samples were placed into an incubator at 40°C with a water pan in the bottom to simulate a relative humidity greater than 95%. The 4th sample replicate (baseline) was stored at room temperature and placed in a plastic specimen pouch after 3 hours of drying.

All samples that remained in a humid environment demonstrated total categorical agreement of 95.0%, with increasing mean biases of −13.3% (1 hour), −22.2% (3 hours) and −44.6% (22 hours, Table 16). Samples that did not have categorical agreement with baseline results had baseline concentrations within 11% of the DBS clinical cutoff (0.185 U/mL). These results indicate that prolonged drying (> 1 hour) in a humid environment may lead to unacceptable negative bias in sample results. Risk of donors drying samples inappropriately can be mitigated by appropriate design of donor materials including instructions for use (IFUs) and demonstration videos.

**Table 16.** Impact of humidity during drying for DBS samples for 1, 3 and 22 hours).

| Sample | Drying in Humid Environment Time Prior to Storage (Hrs) |  |  |  |  |  |  |  |  |  |  |  |  |  |
| --- | --- | --- | --- | --- | --- | --- | --- | --- | --- | --- | --- | --- | --- | --- |
|  | Baseline | (3 Hrs) | 1 Hr |  |  |  | 3 Hrs |  |  |  | 22 Hrs |  |  |  |
|  | U/mL | Result | U/mL | Bias | Result | Agree/<br>Disagree | U/mL | Bias | Result | Agree/<br>Disagree | U/mL | Bias | Result | Agree/<br>Disagree |
| 1 | 0.0380 | N | 0.0543 | NA | N | A | 0.0386 | NA | N | A | 0.137 | NA | N | A |
| 2 | 0.0439 | N | 0.0521 | NA | N | A | 0.0384 | NA | N | A | 0.0397 | NA | N | A |
| 3 | 0.0465 | N | 0.0936 | NA | N | A | 0.0632 | NA | N | A | 0.0768 | NA | N | A |
| 4 | 0.0512 | N | 0.0609 | NA | N | A | 0.0259 | NA | N | A | 0.0387 | NA | N | A |
| 5 | 0.0803 | N | 0.0375 | NA | N | A | 0.0330 | NA | N | A | 0.0806 | NA | N | A |
| 6 | 0.166 | N | 0.550 | NA | P | D | 0.175 | NA | N | A | 0.115 | NA | N | A |
| 7 | 0.176 | N | 0.165 | NA | N | A | 0.160 | NA | N | A | 0.0906 | NA | N | A |
| 8 | 0.193 | P | 0.186 | -3.6% | P | A | 0.151 | NA | N | D | 0.0814 | NA | N | D |
| 9 | 0.361 | P | 0.309 | -14.4% | P | A | 0.297 | -17.7% | P | A | 0.192 | -46.8% | P | A |
| 10 | 0.410 | P | 0.368 | -10.2% | P | A | 0.303 | -26.1% | P | A | 0.231 | -43.7% | P | A |
| 11 | 0.488 | P | 0.427 | -12.5% | P | A | 0.370 | -24.2% | P | A | 0.309 | -36.7% | P | A |
| 12 | 0.657 | P | 0.532 | -19.0% | P | A | 0.491 | -25.3% | P | A | 0.333 | -49.3% | P | A |
| 13 | 0.684 | P | 0.593 | -13.3% | P | A | 0.494 | -27.8% | P | A | 0.353 | -48.4% | P | A |
| 14 | 0.837 | P | 0.739 | -11.7% | P | A | 0.684 | -18.3% | P | A | 0.419 | -49.9% | P | A |
| 15 | 0.926 | P | 0.851 | -8.1% | P | A | 0.758 | -18.1% | P | A | 0.530 | -42.8% | P | A |
| 16 | 0.936 | P | 0.789 | -15.7% | P | A | 0.781 | -16.6% | P | A | 0.541 | -42.2% | P | A |
| 17 | 0.987 | P | 0.793 | -19.7% | P | A | 0.690 | -30.1% | P | A | 0.527 | -46.6% | P | A |
| 18 | 1.36 | P | 1.23 | -9.6% | P | A | 1.15 | -15.4% | P | A | 0.791 | -41.8% | P | A |
| 19 | 1.48 | P | 1.23 | -16.9% | P | A | 1.19 | -19.6% | P | A | 0.908 | -38.6% | P | A |
| 20 | 1.52 | P | 1.25 | -17.8% | P | A | 1.10 | -27.6% | P | A | 0.793 | -47.8% | P | A |
|  |  |  | Mean Bias | -13.3% | Total Agreement: | 95.0% | Mean Bias | -22.2% | Total Agreement: | 95.0% | Mean Bias | -44.6% | Total Agreement: | 95.0% |

N – negative, P – positive, A – results agree, D – results disagree

#### 9.3. Effects of Alcohol Exposure to DBS cards

The potential effects of alcohol exposure to DBS cards prior to addition of blood was investigated in the event that an individual did not allow their finger to dry after sterilization with an alcohol wipe. This study was performed by wiping an alcohol pad on a DBS card immediately prior to the addition of contrived blood samples to the card. For this study all samples were stored after three hours of drying and stored at room temperature. All samples demonstrated total categorical agreement of 100%, with a mean bias of −11.0% (Table 17). As this study represents a worst case scenario to alcohol exposure (saturation of DBS card with alcohol), these results are considered acceptable. Risk of donors exposing alcohol to blood can be mitigated by appropriate design of donor materials including instructions for use (IFUs) and demonstration videos.

**Table 17.** DBS alcohol exposure results.

| Sample | Baseline |  | Alcohol |  |  |  |
| --- | --- | --- | --- | --- | --- | --- |
|  | U/mL | Result | U/mL | Bias | Result | Agree/<br>Disagree |
| 1 | 0.0380 | N | 0.138 | NA | N | A |
| 2 | 0.0439 | N | 0.0727 | NA | N | A |
| 3 | 0.0465 | N | 0.0943 | NA | N | A |
| 4 | 0.0512 | N | 0.0558 | NA | N | A |
| 5 | 0.0803 | N | 0.182 | NA | N | A |
| 6 | 0.166 | N | 0.174 | NA | N | A |
| 7 | 0.176 | N | 0.139 | NA | N | A |
| 8 | 0.193 | P | 0.220 | 14.0% | P | A |
| 9 | 0.361 | P | 0.360 | -0.3% | P | A |
| 10 | 0.410 | P | 0.348 | -15.1% | P | A |
| 11 | 0.488 | P | 0.411 | -15.8% | P | A |
| 12 | 0.657 | P | 0.431 | -34.4% | P | A |
| 13 | 0.684 | P | 0.833 | 21.8% | P | A |
| 14 | 0.837 | P | 0.754 | -9.9% | P | A |
| 15 | 0.926 | P | 0.855 | -7.7% | P | A |
| 16 | 0.936 | P | 0.812 | -13.2% | P | A |
| 17 | 0.987 | P | 0.808 | -18.1% | P | A |
| 18 | 1.36 | P | 1.11 | -18.4% | P | A |
| 19 | 1.48 | P | 1.15 | -22.3% | P | A |
| 20 | 1.52 | P | 1.17 | -23.0% | P | A |
|  |  |  | Mean Bias | -11.0% | Total<br>Agreement: | 100.0% |

N – negative, P – positive, A – results agree, D – results disagree

#### 9.4. Effects of Finger Contamination

The effects of an unsterilized finger being exposed to a DBS spot was investigated by pressing an ungloved finger directly on a DBS card immediately prior to the addition of contrived blood samples. For this study all samples were stored after three hours of drying and stored at room temperature. Total qualitative categorical agreement of 95.0% was observed with mean bias of 1.3% (Table 18). As this study represents a worst case scenario to finger exposure, these results are considered acceptable. The one sample that did not have categorical agreement with baseline results had a baseline concentration of 0.208 U/mL which is within 13% of the DBS clinical cutoff (0.185 U/mL). Risk of donors contaminating DBS cards prior to addition of blood can be mitigated by appropriate design of donor materials including instructions for use (IFUs) and demonstration videos.

**Table 18.** DBS spot contamination with an unsterilized finger results.

| Sample | Baseline |  | Finger |  |  |  |
| --- | --- | --- | --- | --- | --- | --- |
|  | U/mL | Result | U/mL | Bias | Result | Agree/<br>Disagree |
| 1 | 0.0342 | N | 0.0416 | NA | N | A |
| 2 | 0.0364 | N | 0.0444 | NA | N | A |
| 3 | 0.0382 | N | 0.0524 | NA | N | A |
| 4 | 0.0402 | N | 0.0372 | NA | N | A |
| 5 | 0.0450 | N | 0.0557 | NA | N | A |
| 6 | 0.208 | P | 0.175 | NA | N | D |
| 7 | 0.258 | P | 0.288 | 11.6% | P | A |
| 8 | 0.266 | P | 0.270 | 1.5% | P | A |
| 9 | 0.407 | P | 0.400 | -1.7% | P | A |
| 10 | 0.443 | P | 0.410 | -7.4% | P | A |
| 11 | 0.458 | P | 0.479 | 4.6% | P | A |
| 12 | 0.580 | P | 0.568 | -2.1% | P | A |
| 13 | 0.591 | P | 0.592 | 0.2% | P | A |
| 14 | 0.595 | P | 0.606 | 1.8% | P | A |
| 15 | 0.652 | P | 0.712 | 9.2% | P | A |
| 16 | 0.853 | P | 0.829 | -2.8% | P | A |
| 17 | 0.914 | P | 0.755 | -17.4% | P | A |
| 18 | 0.969 | P | 1.03 | 6.3% | P | A |
| 19 | 1.03 | P | 1.11 | 7.8% | P | A |
| 20 | 1.33 | P | 1.42 | 6.8% | P | A |
|  |  |  | Total Bias: | 1.3% | Total Agreement: | 95.0% |

N – negative, P – positive, A – results agree, D – results disagree

### 10. Analytic Interference determination for DBS samples

Studies were also performed to test the effects of different interferences on the measurement of SARS-CoV-2 antibodies from DBS samples. Contrived blood samples were prepared with antibody concentrations ranging from 0.0342 U/mL (∼0.2 times DBS cutoff) to 1.52 U/mL (∼175 times DBS cutoff); most samples were within 5 times the DBS cutoff. All samples used to generate baseline results were spotted and dried for three hours prior to being stored. For acceptance of results, biases were determined to baseline measurements for results greater than the assay’s LOD. A mean bias of 20.0% was used as quantitative acceptance following FDA guidance for ligand binding assays (*3*). For acceptance of qualitative results, a total categorical agreement of 95.0% was utilized based on guidance from the FDA’s Home Specimen Collection Serology Template (*5*).

#### 10.1. DBS Hemolysis Interference Study

To test the effects of hemolysis, contrived liquid blood samples were prepared in duplicate aliquots. One of the aliquots was frozen at −70°C for at least 30 minutes and thawed in order to lyse the red blood cells. Both aliquots (lysed and un-lysed) were then spotted onto DBS cards, dried and stored prior to testing. A total qualitative categorical agreement of 95.0% to un-lysed measurements was observed, one sample disagreed (D) with baseline results with a measured concentration of 0.208 U/mL, within 13% of the DBS clinical cutoff (0.185 U/mL). Lysed DBS samples demonstrated an acceptable mean bias of 3.6% compared to baseline measurements (Table 19).

**Table 19.** DBS hemolysis interference results.

| Hemolysis Interference DBS results (U/mL) |  |  |  |  |  |  |
| --- | --- | --- | --- | --- | --- | --- |
| Sample | Baseline |  | Hemolyzed |  |  |  |
|  | U/mL | Result | U/mL | Bias | Result | Agree/<br>Disagree |
| 1 | 0.0342 | N | 0.0692 | NA | N | A |
| 2 | 0.0364 | N | 0.0774 | NA | N | A |
| 3 | 0.0382 | N | 0.0516 | NA | N | A |
| 4 | 0.0402 | N | 0.0658 | NA | N | A |
| 5 | 0.045 | N | 0.0538 | NA | N | A |
| 6 | 0.208 | P | 0.184 | -11.5% | N | D |
| 7 | 0.258 | P | 0.319 | 23.6% | P | A |
| 8 | 0.266 | P | 0.291 | 9.4% | P | A |
| 9 | 0.407 | P | 0.443 | 8.8% | P | A |
| 10 | 0.443 | P | 0.432 | -2.5% | P | A |
| 11 | 0.458 | P | 0.509 | 11.1% | P | A |
| 12 | 0.580 | P | 0.586 | 1.0% | P | A |
| 13 | 0.591 | P | 0.692 | 17.1% | P | A |
| 14 | 0.595 | P | 0.661 | 11.1% | P | A |
| 15 | 0.652 | P | 0.677 | 3.8% | P | A |
| 16 | 0.853 | P | 0.803 | -5.9% | P | A |
| 17 | 0.914 | P | 0.854 | -6.6% | P | A |
| 18 | 0.969 | P | 0.965 | -0.4% | P | A |
| 19 | 1.03 | P | 1.07 | 3.9% | P | A |
| 20 | 1.33 | P | 1.22 | -8.3% | P | A |
|  |  |  | Total Bias: | 3.6% | Total Agreement: | 95.0% |

N – negative, P – positive, A – results agree, D – results disagree

#### 10.2. Interferences from Endogenous Substances

Endogenous interferents were spiked directly into serum samples prior to adding red blood cells to make contrived blood samples, then spotted onto DBS cards (Table 20). To account for dilution of the serum with the interferent a paired (un-spiked) serum was diluted with a confirmed negative serum (blank) using the same dilution ratio. The spiked and un-spiked serum were then made into contrived blood samples and spotted onto DBS cards. All samples followed the recommended drying and overnight storage, samples were extracted the following day and results were compared to paired samples without interferents. For acceptance criteria of results, the FDA guidelines were used for quantitative and qualitative analysis (*3,5*).

**Table 20.** Endogenous interferent spiking scheme.

| Sun Diagnostics Interferent (Catalog #) | Test Concentration | Interferent Concentration | Serum Relative Volume | Interferent Relative Volume |
| --- | --- | --- | --- | --- |
| Triglycerides (INT-01T) | 3000 mg/dL | >15,000 mg/dL (5X) | 4 parts | 1 part |
| Total Protein (INT-01P) | 12 g/dL* | >20 g/dL (2X) | 4 parts | 3 parts |
| Bilirubin, conjugated (INT-01B) | 20 mg/dL | >400 mg/dL (20X) | 19 parts | 1 part |
| Bilirubin, unconjugated (INT-01BU) | 20 mg/dL | >400 mg/dL (20X) | 19 parts | 1 part |
| Biotin (INT-09) | 3,510 ng/mL | 70,200 ng/mL (20X) | 19 parts | 1 part |

\*Assuming a serum concentration of 6 g/dL.

#### 10.3. DBS Triglyceride Interference Study

When spiking samples with triglycerides at a concentration of 3000 mg/dL, DBS results were found to have a categorical agreement of 96.7%. The one sample that did not have categorical agreement had a baseline concentration of 0.179 U/mL which is within 4.0% of the DBS clinical cutoff (0.185 U/mL). Overall, the triglyceride spiked DBS samples had a mean bias of 4.5% compared to baseline results (Table 21). These results were acceptable under the FDA guidelines for the mean bias and categorical agreement (*3,5*).

**Table 21.** DBS triglycerides interference results.

| Sample | Baseline |  | Triglyceride Interference (3000 mg/dL) |  |  |  |
| --- | --- | --- | --- | --- | --- | --- |
|  | U/mL | Result | U/mL | Bias | Result | Agree/<br>Disagree |
| 1 | 0.0512 | N | 0.0411 | NA | N | A |
| 2 | 0.0557 | N | 0.0676 | NA | N | A |
| 3 | 0.0595 | N | 0.0483 | NA | N | A |
| 4 | 0.0709 | N | 0.0527 | NA | N | A |
| 5 | 0.0733 | N | 0.0825 | NA | N | A |
| 6 | 0.0747 | N | 0.0411 | NA | N | A |
| 7 | 0.0791 | N | 0.144 | NA | N | A |
| 8 | 0.0797 | N | 0.0731 | NA | N | A |
| 9 | 0.0809 | N | 0.0703 | NA | N | A |
| 10 | 0.0835 | N | 0.073 | NA | N | A |
| 11 | 0.136 | N | 0.121 | NA | N | A |
| 12 | 0.179 | N | 0.218 | NA | P | D |
| 13 | 0.21 | P | 0.228 | 8.6% | P | A |
| 14 | 0.296 | P | 0.380 | 28.4% | P | A |
| 15 | 0.314 | P | 0.328 | 4.5% | P | A |
| 16 | 0.389 | P | 0.386 | -0.8% | P | A |
| 17 | 0.459 | P | 0.392 | -14.6% | P | A |
| 18 | 0.461 | P | 0.363 | -21.3% | P | A |
| 19 | 0.732 | P | 0.786 | 7.4% | P | A |
| 20 | 0.768 | P | 0.797 | NA | P | A |
| 21 | 0.86 | P | 0.876 | 1.5% | P | A |
| 22 | 0.861 | P | 1.21 | 40.5% | P | A |
| 23 | 0.945 | P | 0.860 | -9.0% | P | A |
| 24 | 0.96 | P | 0.797 | -17.0% | P | A |
| 25 | 1.01 | P | 1.19 | 17.8% | P | A |
| 26 | 1.02 | P | 0.999 | -2.1% | P | A |
| 27 | 1.05 | P | 1.08 | 2.9% | P | A |
| 28 | 1.1 | P | 1.09 | -0.9% | P | A |
| 29 | 1.53 | P | 1.72 | 12.4% | P | A |
| 30 | 1.60 | P | 1.90 | 18.8% | P | A |
| Mean Bias: |  |  |  | 4.5% | Agreement: | 96.7% |

N – negative, P – positive, A – results agree, D – results disagree

#### 10.4. DBS Total Protein Interference Study

Following spiking of DBS samples to create additive protein concentrations of 12 g/dL, measurements indicated a total qualitative categorical agreement of 100% and mean bias of 17.7% when compared to baseline measurements (Table 22). These results were acceptable under the FDA criteria for the mean bias and categorical agreement (*3,5*).

**Table 22.** DBS total protein interference results.

| Sample | Baseline |  | Protein Interference (12 g/dL) |  |  |  |
| --- | --- | --- | --- | --- | --- | --- |
|  | U/mL | Result | U/mL | Bias | Result | Agree/<br>Disagree |
| 1 | 0.0532 | N | 0.0667 | NA | N | A |
| 2 | 0.0567 | N | 0.0834 | NA | N | A |
| 3 | 0.0572 | N | 0.0973 | NA | N | A |
| 4 | 0.0629 | N | 0.112 | NA | N | A |
| 5 | 0.0635 | N | 0.0995 | NA | N | A |
| 6 | 0.0675 | N | 0.144 | NA | N | A |
| 7 | 0.0678 | N | 0.0894 | NA | N | A |
| 8 | 0.0690 | N | 0.110 | NA | N | A |
| 9 | 0.0778 | N | 0.0618 | NA | N | A |
| 10 | 0.0818 | N | 0.0969 | NA | N | A |
| 11 | 0.130 | N | 0.159 | NA | N | A |
| 12 | 0.240 | P | 0.281 | 17.1% | P | A |
| 13 | 0.252 | P | 0.340 | 34.9% | P | A |
| 14 | 0.265 | P | 0.326 | 23.0% | P | A |
| 15 | 0.268 | P | 0.355 | 32.5% | P | A |
| 16 | 0.289 | P | 0.278 | -3.8% | P | A |
| 17 | 0.291 | P | 0.369 | 26.8% | P | A |
| 18 | 0.313 | P | 0.377 | 20.4% | P | A |
| 19 | 0.317 | P | 0.370 | 16.7% | P | A |
| 20 | 0.328 | P | 0.363 | 10.7% | P | A |
| 21 | 0.336 | P | 0.418 | 24.4% | P | A |
| 22 | 0.386 | P | 0.455 | 17.9% | P | A |
| 23 | 0.395 | P | 0.404 | 2.3% | P | A |
| 24 | 0.580 | P | 0.582 | 0.3% | P | A |
| 25 | 0.604 | P | 0.816 | 35.1% | P | A |
| 26 | 0.625 | P | 0.644 | 3.0% | P | A |
| 27 | 0.653 | P | 0.609 | -6.7% | P | A |
| 28 | 0.685 | P | 0.877 | 28.0% | P | A |
| 29 | 0.714 | P | 1.03 | 44.3% | P | A |
| 30 | 1.07 | P | 1.18 | 10.3% | P | A |
| Mean Bias: |  |  |  | 17.7% | Agreement: | 100.0% |

N – negative, P – positive, A – results agree, D – results disagree

#### 10.5. DBS Bilirubin Interference Studies

Spiking of conjugated bilirubin (up to 20 mg/dL) into DBS samples demonstrated a categorical agreement of 96.7% with baseline DBS samples with a mean bias of −2.8% (Table 23). The one sample that did not have categorical agreement had a baseline concentration of 0.0653 U/mL and a spiked concentration of 0.215 U/mL which is within 16.0% of the DBS clinical cutoff (0.185 U/mL). Spiking of DBS samples with unconjugated bilirubin demonstrated a categorical agreement of 100.0% with baseline results with a mean bias of −8.7% (Table 23). These results were acceptable under the FDA guidance for mean bias and categorical agreement (*3,5*).

**Table 23.** DBS conjugated and unconjugated bilirubin interference results.

| Sample | Baseline |  | Conjugated Bilirubin Results |  |  |  | Unconjugated Bilirubin Results |  |  |  |
| --- | --- | --- | --- | --- | --- | --- | --- | --- | --- | --- |
|  | U/mL | Result | U/mL | Bias | Result | Agree/<br>Disagree | U/mL | Bias | Result | Agree/<br>Disagree |
| 1 | 0.0653 | N | 0.215 | NA | P | D | 0.140 | NA | N | A |
| 2 | 0.0781 | N | 0.151 | NA | N | A | 0.0941 | NA | N | A |
| 3 | 0.0873 | N | 0.106 | NA | N | A | 0.0939 | NA | N | A |
| 4 | 0.0950 | N | 0.122 | NA | N | A | 0.0880 | NA | N | A |
| 5 | 0.101 | N | 0.134 | NA | N | A | 0.0871 | NA | N | A |
| 6 | 0.103 | N | 0.0707 | NA | N | A | 0.0662 | NA | N | A |
| 7 | 0.105 | N | 0.0886 | NA | N | A | 0.0671 | NA | N | A |
| 8 | 0.109 | N | 0.0901 | NA | N | A | 0.0697 | NA | N | A |
| 9 | 0.114 | N | 0.0788 | NA | N | A | 0.0637 | NA | N | A |
| 10 | 0.132 | N | 0.134 | NA | N | A | 0.0798 | NA | N | A |
| 11 | 0.283 | P | 0.248 | -12.4% | P | A | 0.251 | -11.3% | P | A |
| 12 | 0.326 | P | 0.343 | 5.2% | P | A | 0.352 | 8.0% | P | A |
| 13 | 0.429 | P | 0.430 | 0.2% | P | A | 0.362 | -15.6% | P | A |
| 14 | 0.449 | P | 0.412 | -8.2% | P | A | 0.365 | -18.7% | P | A |
| 15 | 0.453 | P | 0.431 | -4.9% | P | A | 0.454 | 0.2% | P | A |
| 16 | 0.470 | P | 0.426 | -9.4% | P | A | 0.399 | -15.1% | P | A |
| 17 | 0.472 | P | 0.509 | 7.8% | P | A | 0.499 | 5.7% | P | A |
| 18 | 0.490 | P | 0.521 | 6.3% | P | A | 0.542 | 10.6% | P | A |
| 19 | 0.495 | P | 0.444 | -10.3% | P | A | 0.387 | -21.8% | P | A |
| 20 | 0.558 | P | 0.594 | 6.5% | P | A | 0.409 | -26.7% | P | A |
| 21 | 1.11 | P | 1.02 | -8.1% | P | A | 0.941 | -15.2% | P | A |
| 22 | 1.13 | P | 1.05 | -7.1% | P | A | 1.17 | 3.5% | P | A |
| 23 | 1.15 | P | 1.10 | -4.3% | P | A | 0.985 | -14.3% | P | A |
| 24 | 1.19 | P | 1.24 | 4.2% | P | A | 1.17 | -1.7% | P | A |
| 25 | 1.32 | P | 1.32 | 0.0% | P | A | 1.19 | -9.8% | P | A |
| 26 | 1.35 | P | 1.31 | -3.0% | P | A | 1.11 | -17.8% | P | A |
| 27 | 1.42 | P | 1.33 | -6.3% | P | A | 1.20 | -15.5% | P | A |
| 28 | 1.47 | P | 1.35 | -8.2% | P | A | 1.23 | -16.3% | P | A |
| 29 | 1.59 | P | 1.42 | -10.7% | P | A | 1.50 | -5.0% | P | A |
| 30 | 2.10 | P | 2.26 | 7.6% | P | A | 2.19 | 4.3% | P | A |
|  |  |  | Mean Bias: | -2.8% | Agreement: | 96.7% | Mean Bias: | -8.7% | Agreement: | 100.0% |

N – negative, P – positive, A – results agree, D – results disagree

#### 10.6. Interferences from Biotin on DBS

Biotin interference was tested by spiking DBS samples to a biotin concentration of 3,510 ng/mL. A total categorical agreement of 96.6% with baseline DBS samples was observed (Table 24). The one sample that did not agree with baseline measurements had a measured baseline concentration (0.205 U/mL) within 11% of the DBS clinical cutoff (0.185 U/mL). These results indicate that DBS measurements in the presence of biotin levels up to 3,510 ng/mL are acceptable under the FDA guidance for the mean bias and categorical agreement (*3,5*).

**Tables S24.** DBS biotin interference results.

**Tables 24. DBS biotin interference results**
| Sample | Baseline |  | Biotin (3510ng/mL) |  |  |  |
| --- | --- | --- | --- | --- | --- | --- |
|  | U/mL | Result | U/mL | Bias | Result | Agree/<br>Disagree |
| 1 | 0.112 | N | 0.123 | NA | N | A |
| 2 | 0.125 | N | 0.0852 | NA | N | A |
| 3 | 0.128 | N | 0.0859 | NA | N | A |
| 4 | 0.149 | N | 0.0766 | NA | N | A |
| 5 | 0.154 | N | 0.0958 | NA | N | A |
| 6 | 0.170 | N | 0.0929 | NA | N | A |
| 7 | 0.182 | N | 0.103 | NA | N | A |
| 8 | 0.205 | P | 0.101 | NA | N | D |
| 9 | 0.245 | P | 0.199 | -18.8% | P | A |
| 10 | 0.324 | P | 0.344 | 6.2% | P | A |
| 11 | 0.328 | P | 0.215 | -34.5% | P | A |
| 12 | 0.347 | P | 0.332 | -4.3% | P | A |
| 13 | 0.352 | P | 0.302 | -14.2% | P | A |
| 14 | 0.397 | P | 0.337 | -15.1% | P | A |
| 15 | 0.511 | P | 0.502 | -1.8% | P | A |
| 16 | 0.575 | P | 0.596 | 3.7% | P | A |
| 17 | 0.643 | P | 0.648 | 0.8% | P | A |
| 18 | 0.700 | P | 0.837 | 19.6% | P | A |
| 19 | 0.710 | P | 0.798 | 12.4% | P | A |
| 20 | 0.938 | P | 1.04 | 10.9% | P | A |
| 21 | 1.07 | P | 0.989 | NA | P | A |
| 22 | 1.16 | P | 1.33 | 14.7% | P | A |
| 23 | 1.17 | P | 1.36 | 16.2% | P | A |
| 24 | 1.26 | P | 1.46 | 15.9% | P | A |
| 25 | 1.30 | P | 1.39 | 6.9% | P | A |
| 26 | 1.34 | P | 1.49 | 11.2% | P | A |
| 27 | 1.37 | P | 1.64 | 19.7% | P | A |
| 28 | 4.21 | P | 4.73 | 12.4% | P | A |
| 29 | 7.63 | P | 7.93 | 3.9% | P | A |
| Mean Bias: |  |  |  | 3.3% | Agreement: | 96.6% |

N – negative, P – positive, A – results agree, D – results disagree

#### 10.7. Interferences from Pharmaceutical Substances on DBS Analysis

Serum samples were spiked with pharmaceutical interferents (from Cerilliant Analytical Reference Standards) and then red blood cells were added to make contrived blood samples that were spotted on DBS cards (Table 25). To account for dilution of the serum with the interferent a paired (un-spiked) serum was diluted with a confirmed negative serum (blank) using the same dilution ratio. The spiked and un-spiked serum were then made into contrived blood samples and spotted onto DBS cards. Samples dried for the recommended time of three hours prior to storage.

**Table 25.** Pharmaceutical substance interference spiking scheme.

| Cerilliant Interference Mix (Catalog #) | Pharmaceutical Substance | Test Concentration (µg/mL) | Interferent Concentration (µg/mL) | Serum Relative Volume | Interferent Relative Volume |
| --- | --- | --- | --- | --- | --- |
| 1<br>(I-023) | (-)-Cotinine | 100 | 1000 | 9 parts | 1 part |
|  | (-)-Nicotine | 100 | 1000 |  |  |
|  | Acetaminophen | 100 | 1000 |  |  |
|  | Caffeine | 100 | 1000 |  |  |
|  | Ibuprofen | 100 | 1000 |  |  |
|  | Naproxen | 100 | 1000 |  |  |
|  | Phentermine | 10 | 100 |  |  |
|  | R,R(-)-Pseudoephedrine, List I | 10 | 100 |  |  |
| 2<br>(I-024) | Gabapentin | 10 | 100 | 9 parts | 1 part |
|  | Pregabalin | 10 | 100 |  |  |
|  | Salicylic acid | 10 | 100 |  |  |
|  | Valproic acid | 10 | 100 |  |  |
|  | Vigabatrin | 10 | 100 |  |  |

DBS samples spiked with Cerilliant mix 1 had a total categorical agreement of 96.6% with baseline DBS results (Table 26). One of the thirty Cerilliant mix 1 DBS samples was in disagreement with baseline; DBS results were less than 1% greater than the DBS clinical cutoff (0.186 versus the cutoff of 0.185 U/mL). Mean bias was 5.8% when compared to baseline results. Samples that were spiked with Cerilliant mix 2 had a total categorical agreement of 100.0% with baseline DBS results. Overall, Cerilliant mix 2DBS results had a mean bias −0.2% when compared to baseline DBS results (Table 27). These results indicate that DBS samples containing supra-physiological levels of tested drugs from Cerilliant mix 1 and 2 are acceptable under the FDA guidance for the mean bias and categorical agreement (*3,5*). Cerilliant mix 1 and 2 DBS results were within the acceptance criteria provided by the FDA.

**Table 26.** DBS Cerilliant Mix 1 interference results.

| Sample | Baseline |  | Cerilliant Mix 1 |  |  |  |
| --- | --- | --- | --- | --- | --- | --- |
|  | U/mL | Result | U/mL | Bias | Result | Agree/<br>Disagree |
| 1 | 0.0575 | N | 0.0771 | NA | N | A |
| 2 | 0.0580 | N | 0.117 | NA | N | A |
| 3 | 0.0646 | N | 0.0959 | NA | N | A |
| 4 | 0.0653 | N | 0.100 | NA | N | A |
| 5 | 0.0660 | N | 0.0769 | NA | N | A |
| 6 | 0.0772 | N | 0.0564 | NA | N | A |
| 7 | 0.0864 | N | 0.0869 | NA | N | A |
| 8 | 0.0910 | N | 0.0727 | NA | N | A |
| 9 | 0.104 | N | 0.0547 | NA | N | A |
| 10 | 0.171 | N | 0.186 | NA | P | D |
| 11 | 0.226 | P | 0.311 | 37.6% | P | A |
| 12 | 0.252 | P | 0.246 | -2.4% | P | A |
| 13 | 0.276 | P | 0.262 | -5.1% | P | A |
| 14 | 0.434 | P | 0.415 | -4.4% | P | A |
| 15 | 0.467 | P | 0.403 | -13.7% | P | A |
| 16 | 0.469 | P | 0.435 | -7.2% | P | A |
| 17 | 0.514 | P | 0.487 | -5.3% | P | A |
| 18 | 0.528 | P | 0.469 | -11.2% | P | A |
| 19 | 0.546 | P | 0.578 | 5.9% | P | A |
| 20 | 0.881 | P | 0.717 | -18.6% | P | A |
| 21 | 0.940 | P | 1.01 | 7.4% | P | A |
| 22 | 1.03 | P | 1.26 | 22.3% | P | A |
| 23 | 1.14 | P | 1.35 | 18.4% | P | A |
| 24 | 1.18 | P | 1.22 | 3.4% | P | A |
| 25 | 1.21 | P | 1.07 | -11.6% | P | A |
| 26 | 1.31 | P | 1.25 | -4.6% | P | A |
| 27 | 1.60 | P | 1.56 | -2.5% | P | A |
| 28 | 1.95 | P | 1.91 | -2.1% | P | A |
| 29 | 3.54 | P | 3.26 | -7.9% | P | A |
|  |  |  | Total Bias: | 5.8 | Total Agreement: | 96.6% |

N – negative, P – positive, A – results agree, D – results disagree

**Table 27.**
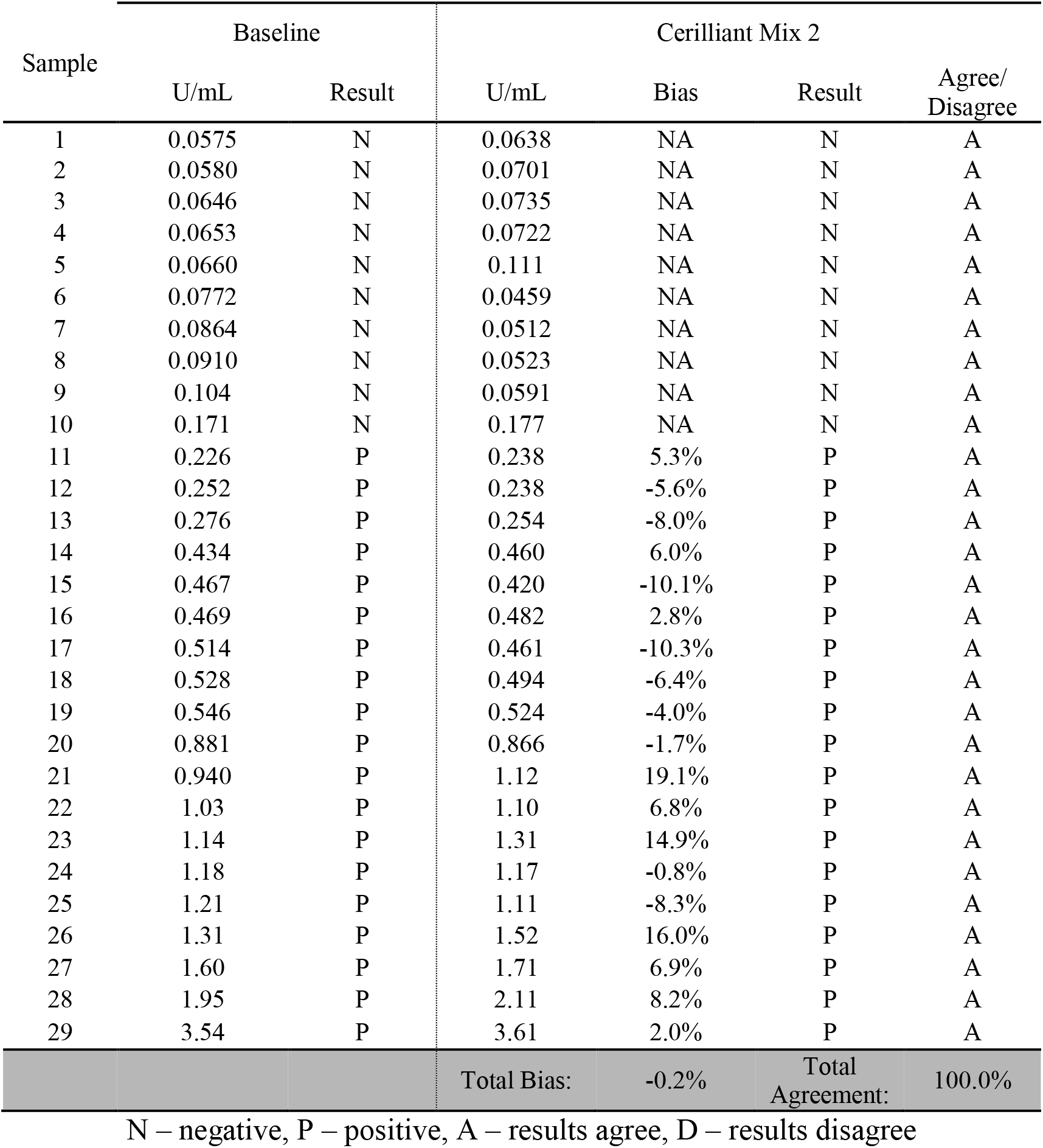
DBS Cerilliant Mix 2 interference results.

| Sample | Baseline |  | Cerilliant Mix 2 |  |  |  |
| --- | --- | --- | --- | --- | --- | --- |
|  | U/mL | Result | U/mL | Bias | Result | Agree/<br>Disagree |
| 1 | 0.0575 | N | 0.0638 | NA | N | A |
| 2 | 0.0580 | N | 0.0701 | NA | N | A |
| 3 | 0.0646 | N | 0.0735 | NA | N | A |
| 4 | 0.0653 | N | 0.0722 | NA | N | A |
| 5 | 0.0660 | N | 0.111 | NA | N | A |
| 6 | 0.0772 | N | 0.0459 | NA | N | A |
| 7 | 0.0864 | N | 0.0512 | NA | N | A |
| 8 | 0.0910 | N | 0.0523 | NA | N | A |
| 9 | 0.104 | N | 0.0591 | NA | N | A |
| 10 | 0.171 | N | 0.177 | NA | N | A |
| 11 | 0.226 | P | 0.238 | 5.3% | P | A |
| 12 | 0.252 | P | 0.238 | -5.6% | P | A |
| 13 | 0.276 | P | 0.254 | -8.0% | P | A |
| 14 | 0.434 | P | 0.460 | 6.0% | P | A |
| 15 | 0.467 | P | 0.420 | -10.1% | P | A |
| 16 | 0.469 | P | 0.482 | 2.8% | P | A |
| 17 | 0.514 | P | 0.461 | -10.3% | P | A |
| 18 | 0.528 | P | 0.494 | -6.4% | P | A |
| 19 | 0.546 | P | 0.524 | -4.0% | P | A |
| 20 | 0.881 | P | 0.866 | -1.7% | P | A |
| 21 | 0.940 | P | 1.12 | 19.1% | P | A |
| 22 | 1.03 | P | 1.10 | 6.8% | P | A |
| 23 | 1.14 | P | 1.31 | 14.9% | P | A |
| 24 | 1.18 | P | 1.17 | -0.8% | P | A |
| 25 | 1.21 | P | 1.11 | -8.3% | P | A |
| 26 | 1.31 | P | 1.52 | 16.0% | P | A |
| 27 | 1.60 | P | 1.71 | 6.9% | P | A |
| 28 | 1.95 | P | 2.11 | 8.2% | P | A |
| 29 | 3.54 | P | 3.61 | 2.0% | P | A |
|  |  |  | Total Bias: | -0.2% | Total Agreement: | 100.0% |

N – negative, P – positive, A – results agree, D – results disagree

## 12. Abbreviations

CLSI: Clinical and Laboratory Standards Institute
CV: Coefficient of variance
DBS: Dried Blood Spot
DF: Degrees of Freedom
EUA: Emergency Use Authorization
FDA: Food and Drug Administration
MS: Mean of Square
n: Number of replicates
N: Negative
NA(or N/A): Not Applicable
NPA: Negative Predictive Agreement
NTP: No test performed
NPV: Negative Predictive Value
P: Positive
PPA: Positive Predictive Agreement
PPV: Positive Predictive Value
QC: Quality control
SD: Standard Deviation
SS: Sum of Squares
U/mL: Units per milliliter

